# Integrated Single-Cell Atlases Reveal an Oral SARS-CoV-2 Infection and Transmission Axis

**DOI:** 10.1101/2020.10.26.20219089

**Authors:** Ni Huang, Paola Perez, Takafumi Kato, Yu Mikami, Kenichi Okuda, Rodney C. Gilmore, Cecilia Domínguez Conde, Billel Gasmi, Sydney Stein, Margaret Beach, Eileen Pelayo, Jose Maldonado, Bernard LaFont, Ricardo Padilla, Valerie Murrah, Robert Maile, Will Lovell, Shannon Wallet, Natalie M. Bowman, Suzanne L Meinig, Matthew C Wolfgang, Saibyasachi N. Choudhury, Mark Novotny, Brian D Aevermann, Richard Scheuermann, Gabrielle Cannon, Carlton Anderson, Julie Marchesan, Mandy Bush, Marcelo Freire, Adam Kimple, Daniel L. Herr, Joseph Rabin, Alison Grazioli, Benjamin N. French, Thomas Pranzatelli, John A. Chiorini, David E. Kleiner, Stefania Pittaluga, Stephen Hewitt, Peter D. Burbelo, Daniel Chertow, NIH COVID-19 Autopsy Consortium, HCA Oral and Craniofacial Biological Network, Karen Frank, Janice Lee, Richard C. Boucher, Sarah A. Teichmann, Blake M. Warner, Kevin M. Byrd

**Affiliations:** Wellcome Sanger Institute, Wellcome Genome Campus, Hinxton, Cambridge CB10 1SA, UK; Salivary Disorders Unit, National Institute of Dental and Craniofacial Research, National Institutes of Health, Bethesda, Maryland, USA; Marsico Lung Institute, University of North Carolina at Chapel Hill, Chapel Hill, NC, USA; Laboratory of Pathology, Center for Cancer Research, National Cancer Institute, National Institutes of Health, Bethesda, Maryland, USA; Emerging Pathogens Section, Department of Critical Care Medicine, NIH Clinical Center, National Institutes of Health, Bethesda, Maryland, USA; AAV Biology Section, National Institute of Dental and Craniofacial Research, National Institutes of Health, Bethesda, Maryland, USA; SARS-CoV-2 Virology Core, Laboratory of Viral Diseases, Division of Intramural Research, NIAID/NIH; Division of Diagnostic Sciences, University of North Carolina Adams School of Dentistry, Chapel Hill, NC, USA; Department of Microbiology & Immunology, University of North Carolina School of Medicine Chapel Hill, NC, USA; Department of Surgery, University of North Carolina at Chapel Hill, Chapel Hill, NC, USA; Division of Oral & Craniofacial Health Sciences, University of North Carolina Adams School of Dentistry, Chapel Hill, NC, USA; Department of Medicine, University of North Carolina at Chapel Hill, Chapel Hill, NC, USA; Department of Genomic Medicine, J. Craig Venter Institute, La Jolla, California, USA; Department of Infectious Disease, J. Craig Venter Institute, La Jolla, California, USA; Department of Informatics, J. Craig Venter Institute, La Jolla, CA, USA; Department of Pathology, University of California San Diego, La Jolla, CA, USA; Center for Gastrointestinal Biology and Disease Advanced Analytics Core, University of North Carolina School of Medicine Chapel Hill, NC, USA; Division of Comprehensive Oral Health, University of North Carolina Adams School of Dentistry, Chapel Hill, NC, USA; ENT, Department of Otolaryngology-Head and Neck Surgery, University of North Carolina School of Medicine Chapel Hill, NC, USA; Department of Shock Trauma Critical Care, University of Maryland School of Medicine, Baltimore, MD, USA; Department of Surgery, R Adams Cowley Shock Trauma Center, University of Maryland School of Medicine, Baltimore, MD, USA; Kidney Diseases Branch, National Institute of Diabetes and Digestive and Kidney Diseases, National Institutes of Health, Bethesda, Maryland, USA; Division of Microbiology, Department of Laboratory Medicine, Clinical Center, National Institutes of Health, Bethesda, Maryland, USA; Craniofacial Anomalies & Regeneration Section, National Institute of Dental and Craniofacial Research, National Institutes of Health, Bethesda, Maryland, USA; Dept Physics, Cavendish Laboratory, JJ Thomson Ave, Cambridge CB3 0HE, UK; ADA Science & Research Institute, Gaithersburg, MD, USA

**Keywords:** SARS-CoV-2, COVID-19, oral cavity, oropharynx, saliva, antibodies, scRNAseq, atlas

## Abstract

Despite signs of infection, the involvement of the oral cavity in COVID-19 is poorly understood. To address this, single-cell RNA sequencing datasets were integrated from human minor salivary glands and gingiva to identify 11 epithelial, 7 mesenchymal, and 15 immune cell clusters. Analysis of SARS-CoV-2 viral entry factor expression showed enrichment in epithelia including the ducts and acini of the salivary glands and the suprabasal cells of the mucosae. COVID-19 autopsy tissues confirmed in vivo SARS-CoV-2 infection in the salivary glands and mucosa. Saliva from SARS-CoV-2-infected individuals harbored epithelial cells exhibiting *ACE2* expression and SARS-CoV-2 RNA. Matched nasopharyngeal and saliva samples found distinct viral shedding dynamics and viral burden in saliva correlated with COVID-19 symptoms including taste loss. Upon recovery, this cohort exhibited salivary antibodies against SARS-CoV-2 proteins. Collectively, the oral cavity represents a robust site for COVID-19 infection and implicates saliva in viral transmission.

## INTRODUCTION

The world remains in the midst of the most serious human pandemic in a century. As of October 15, 2020, Coronavirus Disease 2019 (COVID-19), caused by the novel severe acute respiratory syndrome coronavirus 2 (SARS-CoV-2), has claimed more than one million lives worldwide and critically strained health care infrastructures and the global economy (1, 2). The World Health Organization classifies SARS-CoV-2 as a likely airborne pathogen transmitted by asymptomatic, pre-symptomatic, and symptomatic individuals through close contact via exposure to infected droplets and aerosols (3). While COVID-19 transmission can occur by activities involving the oral cavity, e.g. speaking, breathing, coughing, sneezing, and even singing, most attention has focused on the nasal-lung axis of path of infection (4-9). A recent systematic review of 40 studies including 10,228 COVID-19-infected subjects found oral manifestations in 45% of all cases (14). While oral symptoms such as such taste loss, dry mouth, and mucosal blistering occur in the majority of COVID-19 patients (10-13), little is known about the involvement of the oral cavity SARS-CoV-2 pathogenesis and transmission.

Currently, it remains unknown whether SARS-CoV-2 can infect and replicate widely in tissues of the oral cavity such as the oral mucosae or salivary glands (SG). This information is critical because if these are sites of early infection (15), they could play an important and underappreciated role in transmitting the virus intermucosally to the lungs and/or the gastrointestinal tract via saliva (Figure 1a; 16-18). Saliva may also play a central role in transmitting the virus extraorally via discharged, infectious droplets (19). Saliva is known to harbor SARS-CoV-2 RNA, and is now widely employed in diagnostic testing due to its ease of collection (20-22). Nevertheless, little is known about the source(s) of virus in saliva nor the role of oral virus in oral symptoms or transmission.

**Figure 1.**
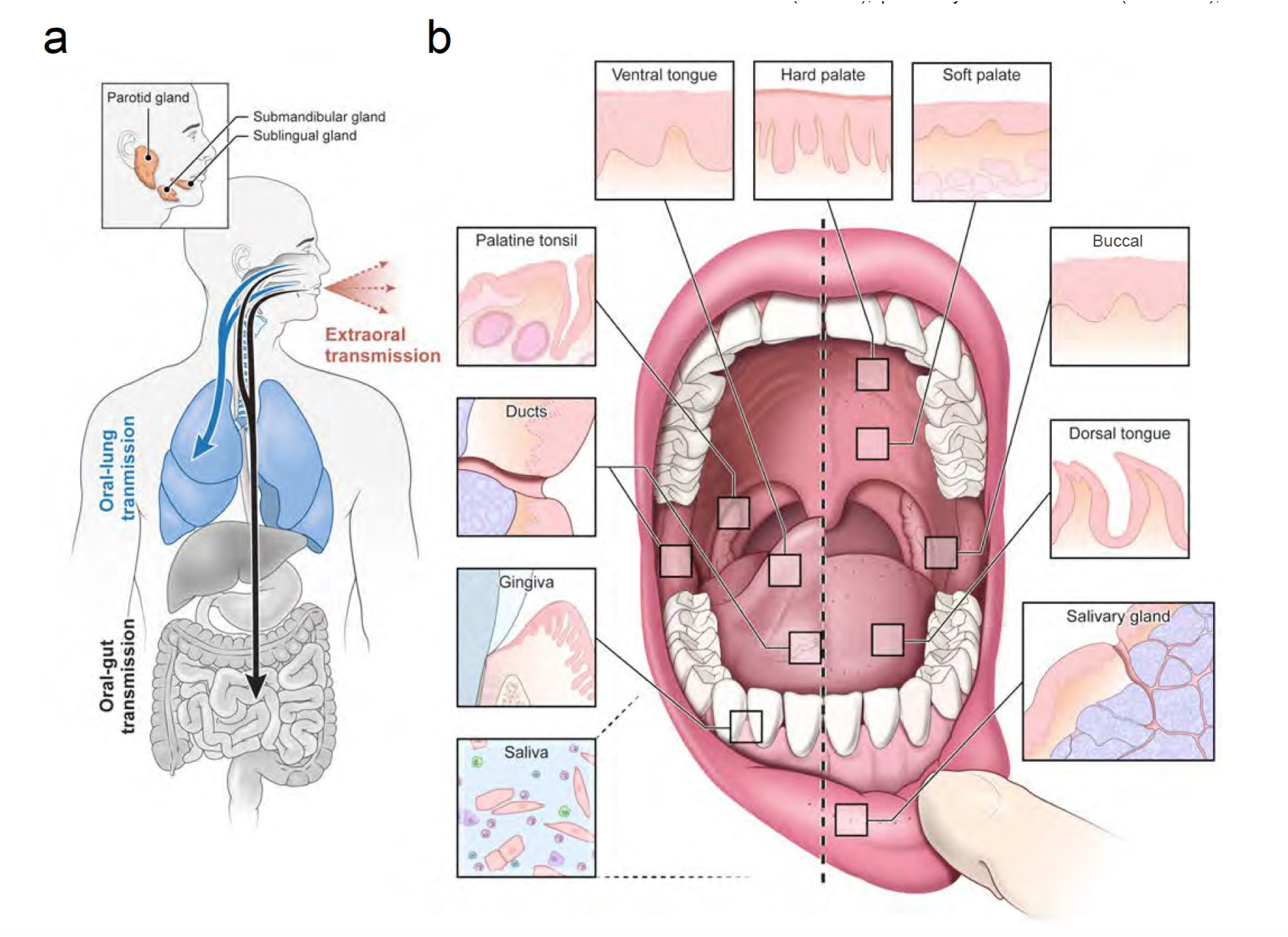
An infection and transmission axis for SARS-CoV-2 among distinct oral niches. **(a)** The contribution of the oral cavity to COVID-19 pathogenesis and transmission has been little explored. It is unknown whether SARS-CoV-2 can infect and replicate in the oral mucosa or glands. This is critical because if the glands or mucosa are sites of early infection, they may play an important and underappreciated role in transmitting virus “intermucosally” to the lungs or gastrointestinal tract. Alternatively, saliva may also play a central role in transmitting the virus extraorally in asymptomatic, pre-symptomatic, or symptomatic individuals. **(b)** The human oral cavity is a diverse collection of tissue niches with potentially unique vulnerabilities to viral infection. These sites include oral mucosae (hard palate, buccal mucosa, dorsal and ventral tongue) as well as the also the terminally differentiated secretory epithelia of the minor saliva glands (distributed in the buccal and labial mucosa, hard and soft palate, ventral and dorsal tongue) and major saliva glands (parotid, submandibular, and sublingual). Nearby are diverse oropharyngeal niches (palatine and lingual tonsils, soft palate). Saliva, a mixture of fluids, electrolytes, proteins, and cells (immune and sloughed mucosal epithelial cells) is made primarily by the saliva glands and empties into the oral cavity where it mixes with other fluids (crevicular fluid) and cells.

SARS-CoV-2 utilizes host entry factors such as ACE2 (23) and TMPRSS family members (TMPRSS2, −4) and understanding the cell types that harbor these receptors is important for determining infection susceptibilities throughout the body (24, 25). The oral cavity and oropharynx are lined by specialized stratified squamous mucosae divided into keratinized (attached gingiva and hard palate) and the non-keratinized (buccal, labial, ventral tongue, oropharyngeal, and unattached gingiva) mucosae. The specialized mucosae of the dorsal and lateral tongue are adapted for taste and contain projections called papillae. Moreover, the entirety of the oral cavity is bathed and protected by saliva. Thus, one of the challenges to understanding the role of the oral cavity in COVID-19 is its heterogeneous niches adapted for feeding, speech, and early digestion (Figure 1b; 26-28). This heterogeneity exists at the regional level and also in the rich cellular diversity in each oral niche (29, 30). While *ACE2* and *TMPRSS2* expression are selectively reported in oral tissues (32-36), there are no comprehensive descriptions of viral entry factor expression to pre-dict vulnerabilities across the oral cavity.

We hypothesize that the diverse epithelia of the oral cavity, from mucosa to SGs, can be infected by SARS-CoV-2 and play a role in both symptomatic and asymptomatic transmission in COVID-19. To test this hypothesis, we generated the first integrated human oral single cell RNA sequencing atlases to predict cellular sources of susceptibilities to SARS-CoV-2 infection. *In situ* hybridization (ISH) was used to confirm *ACE2* and *TMPRSS* expression to epithelia of the SGs and the oral and oropharyngeal mucosae. SARS-CoV-2 infection and replication was confirmed using, autopsy and outpatient samples using ISH and confirmed by polymerase chain reaction (PCR). Nasopharyngeal and saliva samples were studied in a prospective outpatient cohort examining the relationship between saliva viral burden, SARS-CoV-2 RNA in sloughed epithelial cells, and COVID-19 symptoms. Finally, the associations between salivary antibodies, saliva viral load, and clinical status were explored. These data provide a useful approach to predict viral susceptibilities in the oral cavity, demonstrate SARS-CoV-2 infection of oral and oropharyngeal niches, and illustrate the potential for both intermucosal and extraoral transmission of COVID-19 through oral secretions. These data further support recommended public health measures to reduce the risk of transmission of potentially infectious salivary secretions.

## RESULTS

### Human oral tissue atlases reveal unprecedented cell heterogeneity

The SGs and the barrier mucosal epithelia of the oral cavity and oropharynx are likely gateways for viral infection, replication, and transmission. However, considering the remarkable diversity of these tissues, we pre-dicted that SARS-CoV-2 tropism would be nonuniform across oral sites (Figure 1b). To establish the initial database for studying the oral axes in viral pathogenesis, human oral single cell RNA sequencing (scRNAseq) datasets from the SGs (n=5, labial minor) and oral mucosae (n=4, gingiva and hard palate mucosae) were aggregated and analyzed to create a single cell reference atlas of the oral cavity (Figure 2). Utilizing only the minor SG and gingiva, broad host structural and immune cell heterogeneity were identified, including 50 cell types (22 in minor SGs, 28 cell types in mucosa; Supp. Figure 1a) identified. These glandular and mucosal clusters represented unique sets of markers per sample with moderate cell type correlations between cell types (Supp. Figures 1a,b; 2a). Both individual SG and gingival atlases revealed unique and hereunto unreported cell expression signatures per cluster.

**Figure 2.**
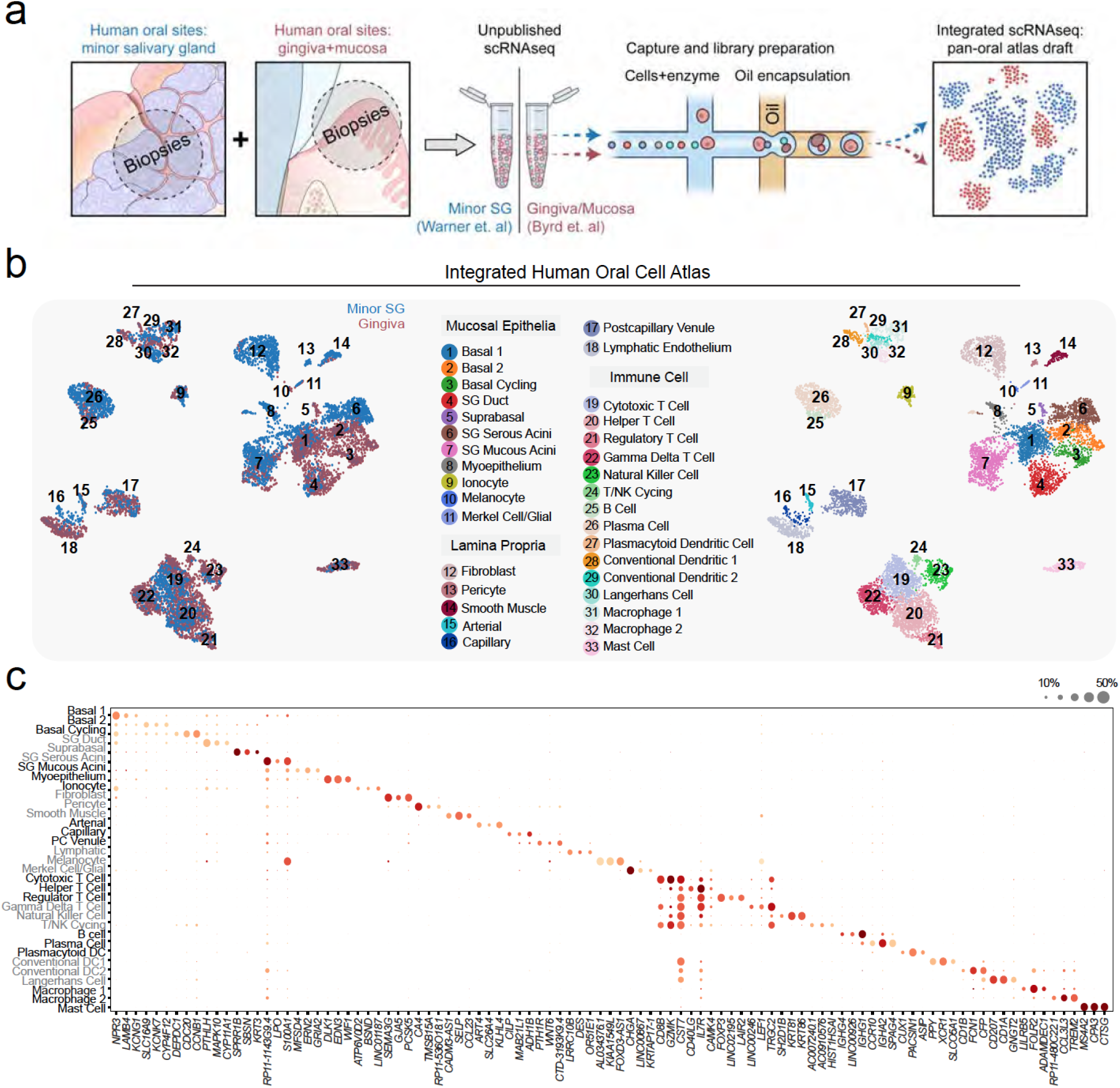
Human pan-oral cell atlas reveals unprecedented cell type heterogeneity. **(a)** To characterize the vulnerabilities of these unique tissues to infection by coronaviruses and other commonly encountered viruses, we integrated unpublished data from human oral gingiva and palatal mucosa (Byrd, 2020) and minor saliva glands (Warner, 2020). To support these findings, we also integrated published data sets from mouse (Supp. Figure 3). **(b)** Overlaid UMAPs delineated by color reveal similar and unique populations based on sample type. (b, Right) These 50 populations were jointly annotated before integration into 33 cell clusters for the first human pan-oral cell atlas. These results illustrate both shared and unique cell populations are represented in the gingival and saliva gland tissues. In total, 33 unique cell types are identified in oral cavity tissues across epithelial, lamina propria, and immune compartments. **(c)** Dot plot expression matrix visualization of the integrated human oral atlas illustrates that cell types are distinguished by unique—and for some oral cell populations—as of yet, undescribed transcriptional signatures.

The minor SGs resemble the submucosal glands of the respiratory system but remain poorly understood in health and disease (37). Minor SGs num-ber ∼1000/person in the human oral cavity and are distributed throughout the most oral mucosae. They secrete a mucous and protein-rich fluid that both lubricates and protect the overlying mucosa. The salivary epithelial compartment of the scRNAseq atlas revealed six major cell types including serous acini (*PRR4, ZG16B*), mucous acini (*MUC5B, BPIFB2*), ducts (*S100A2, KRT14*), myoepithelia (*ACTA2*), glial cells (*CHGA*), and intrigu-ingly ionocytes (*CFTR, FOXI1, V-ATPase*) which were found in higher pro-portion than in the lung (38) and function to regulate ion composition of the ducts (Supp. Figure 1a). The connective tissue compartment consisted of fibroblasts (*DCN, LUM*), endothelium (arterial (*CELP*); venules (*FOXD3*); capillaries (*C4*), smooth muscle (*DES*), and pericytes (*PTH1R*). Reflecting the consistent presence of immune cell, 8 distinct populations of immune cells were identified and was comprised of a high proportion of IgA+ plasma cells, B-lymphocytes (*MS4A1*), T helper (Th) cells (*CD154*), and two subsets of CTLs (*CD8A*), subtype 1 (*EOMES, GZMA*), and subtype 2 (*SPRY, ITGA1*). The myeloid-derived compartment included a subpopula-tion of conventional DC2 (*CD1C*) and two different types of macrophages: subtype 1 (*CD163*) and subtype 2 exhibited an activation signature (*C3, FCGR3A, IFNGR1, CX3CR1*).

The gingiva, along with the periodontal apparatus, supports the teeth and is an indicator of overall dental health status (39), and harbors a diverse immune cell repertoire in health comprised predominately of neutrophils and T cells (40). In this study, the mucosal atlas was generated with mildly inflamed gingival mucosa from healthy individuals and contributed to the identification of 15 immune cell subpopulations (Figure 2b; 30). Five populations of dendritic cells were identified including conventional dendritic cell subtype 1 (*XCR1*) and subtype 2 (*CD300E*); activated dendritic cells (*LAMP3*), plasmacytoid dendritic cells (PACSIN1), and epithelial resident Langerhans cells (*CD207*) (41, 42). Among T cells subtypes, CTLs (*CD8A*), Th (*CD154*), regulatory T cells (*LAIR2*), γδ T cells (*KLRC4*), and mucosal associated invariant T cells (*MAIT; IL23R*); a cycling T/NK cell subset (*POU2AF1*) were cataloged.

Exemplifying the epithelial heterogeneity in the gingiva, 8 distinct popula-tions were identified without overlap with the 5 SG epithelial populations (Figure 2; Supp. Figure 1) in the integrated atlas. Five basal populations (Basal 1-4, and a cycling population), were defined by either expression of *CXCL14, KRT19, SOX6/COL7A1, KRT6A*, and *CCBN1*, (Supp. Figure 1a). It is known that *KRT19* expressing cells defines the non-keratinized gingiva “pocket” epithelium, whereas *KRT6A* marks the basal epithelial population in the keratinized palatal mucosa (43, 44). Suprabasal cells were sparse in this dataset but defined by the KRT6B/KRT16 heterodimer pair and *IVL* (Figure 2; Supp. Figure 1). The fact that signs and symptoms of SARS-CoV-2 infection include oral manifestations raised questions regarding the risk of viral infection and replication in glandular and mucosal epithelial populations, as has been shown with other viruses (31).

### Integrated oral single cell atlases predict SARS-CoV-2 and other viral infections in the oral cavity

To predict viral tropism of SARS-CoV-2, other coronaviruses (SARS-CoV-1, HCoV-NL63, MERS-CoV, HCoV-229E, HCoV-OC43), as well as influenza and rhinovirus, joint annotation of the SG and gingival scRNAseq atlases was performed. Thirty-three unique cell types were identified including 11 epithelial, 7 mesenchymal, and 15 immune cell clusters (Figure 2b,c). The cell-type expression of known viral entry factor genes across cell clusters and virus types (Figure 3a) demonstrated broad susceptibilities to viral infection among the epithelial populations (Figure 3b-c). Similar to SARS-CoV-1 and HCoV-NL63, SARS-CoV-2 spike glycoprotein binds to ACE2 and is reported to be activated by tissue-specific proteases (TMPRSS2, TMPRSS4, TMPRSS11D) and endosomal proteases (CTSB, CTSL, BSG, and possibly, FURIN) to gain entry for viral replication (31).

**Figure 3.**
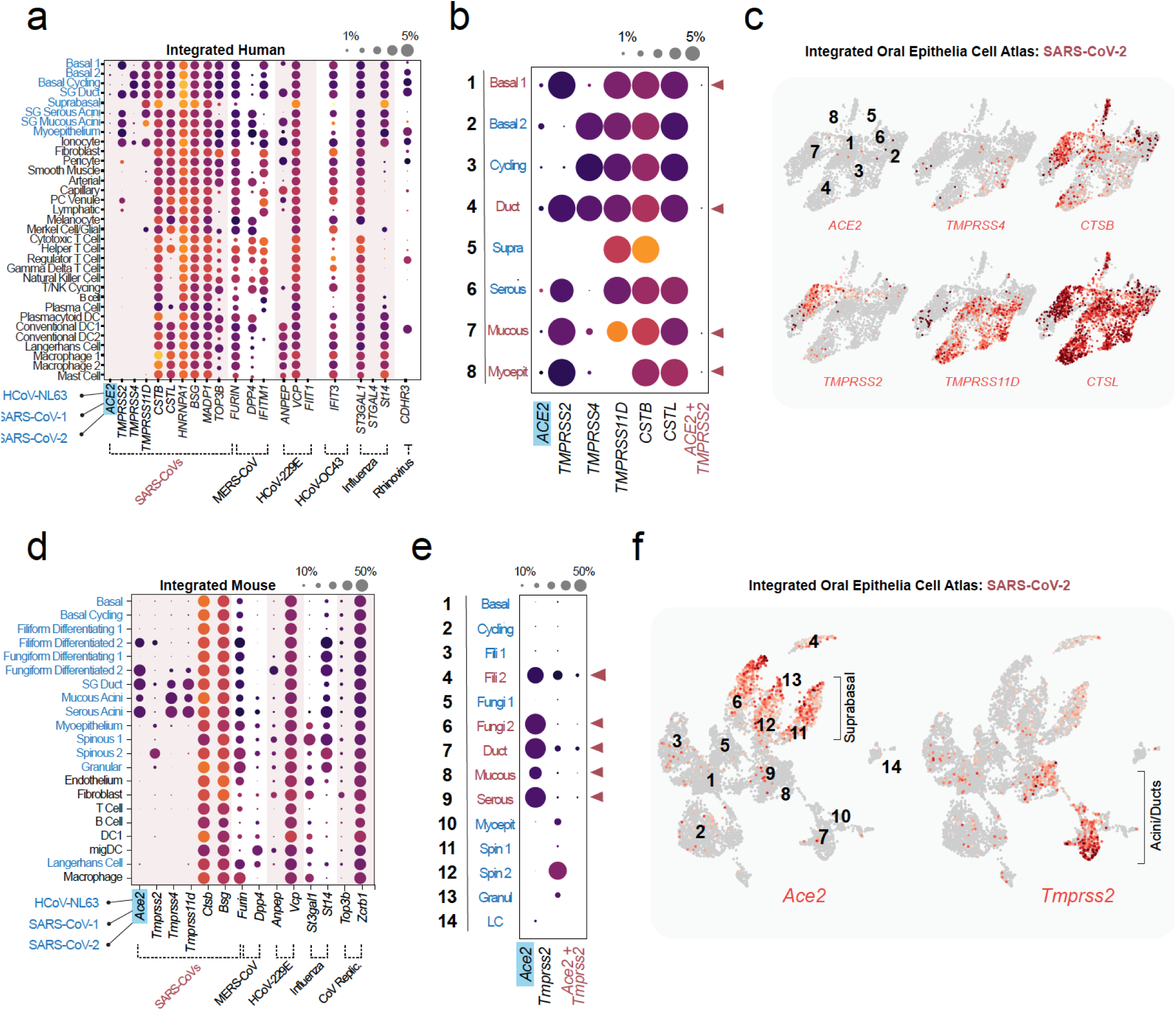
Integrated single-cell atlases uncover broad oral Infection vulnerabilities. **(a-c) (a)** Vulnerabilities to infection by coronaviruses, influenza, and rhinovirus can be predicted based on entry factors expression and visualized using expression matrices. Epithelia appear especially at-risk for viral infection. **(b)** When focused on the 8 epithelial cell populations, vulnerabilities to SARS-CoV-2 were apparent in both glands and mucosa. These results strongly suggest that the oral cavity may be vulnerable to viral infection, especially for SARS-CoV-2. Expression matrices, including low frequency *ACE2/TMPRSS2* co-expressing cells in Basal 1, ducts, mucous acini, and myoepithelial clusters, further support broad SARS-CoV-2 vulnerabilities. **(c)** UMAPS demonstrate distinct cluster vulnerabilities with *ACE2* highest in most oral epithelia; however, expression of proteases demonstrated tissue-specific expression patterns with *TMPRSS2* (enriched for SGs) and *TMPRSS11D* (enriched for mucosal cells). Endosomal proteases, *CTSB* and *CTSL* exhibited broad expression across vulnerable cell types. **(d-f) (d)** Like the human integrated atlas, by using the cell type expression of known host entry factors mouse atlas support the viral vulnerabilities of the lining mucosa and SGs (14 total populations including tissue-resident Langerhans cells; LC) as high risk sites for infection by coronaviruses (SARS-CoVs, MERS, HCoVs), influenza, and rhinovirus. **(e)** These results can be further underscored by looking at co-expression of *Ace2* and *Tmprss2* is restricted to filiform and fungiform differentiated epithelial cells and SG ducts and acini. **(f)** UMAPS of SARS-CoV-2 entry factors demonstrate distinct cluster vulnerabilities with Ace2 highest expressed in suprabasal tissues, Tmprss2 expressed in SGs ducts and acini.

SARS-CoV-2 viral entry factor expression analyses revealed that no single oral epithelia subpopulation appeared at singular risk for infection. *ACE2* expression was detected in 8 oral epithelial clusters including, Basal 1, Basal 2, Basal cycling, SG ducts, suprabasal, and SG serous and mucous acini clusters (Figure 3a-c). These findings support multiple oral epithelial cells are susceptible to SARS-CoV-2 infection. Co-expression of the principal entry factors, *ACE2* and *TMPRSS2*, used to predict tissue susceptibility for SARS-CoV-2 in specific cell types in the nasal cavity, lungs, and gut (4, 25, 45), was infrequent among our epithelial populations likely due to low *ACE2* expression. However, *ACE2* and *TMPRSS2*, co-expression in epithelial cells of the SGs and mucosa, specifically in the SG ducts and acini, and the Basal 1 population of the gingiva (∼0.5% of cells; Figure 3b) was much higher than all other populations. Uniform Manifold Approximation and Projections (UMAPs) of putative SARS-CoV-2 entry factor expression revealed expression of *ACE2* confined primarily to epithelial clusters of the SGs (Figure 3c). Proteases were demonstrated to exhibit tissue-specific expression patterns with *TMPRSS2* enriched in the SG epithelia and TM*PRSS11D* enriched in mucosal cells. Endosomal proteases, *CTSB* and *CTSL*, exhibited broad expression across potentially susceptible cell types.

The generalizability of the human atlas results was extended by including additional oral sites not represented in the human oral single cell atlas by integrating published mouse oral scRNAseq datasets (Supp. Figure 3). Published mouse oral buccal mucosa (46) and dorsal tongue (47), and the major SG, submandibular (47) and parotid (48), datasets were integrated, reannotated, and analyzed for viral entry factor expression (Supp. Figure 3a). Cell type comparison analysis between mouse and human annotations revealed moderate correlation for most clusters (Supp. Figure 2b). Similar to humans, 33 separate, and 21 integrated, cell populations were identified in the mouse oral cavity (Supp. Figure 3b,c), *Ace2* and *Tmprss2/Tmprss4* gene expression was also enriched in mouse oral epithelial clusters (Figure 3d-f). *Ace2* expression was highest in the SGs and differentiated epithelia of the tongue, including co-expression of *Ace2* and *Tmprss2* (Figure 3e,f). Both gland and tongue are sites affected in COVID-19 patients (taste alteration and dry mouth, respectively; 11, 12), and support the likelihood of an oral infection axis in the pathogenesis of SARS-CoV-2.

### SARS-CoV-2 infects multiple sites and can replicate in salivary glands

Employing our integrated atlases to understand the involvement of the oral cavity in COVID-19 pathogenesis, we validated both receptor (*ACE2*) and protease (*TMPRSS2, TMPRSS4*, and *TMPRSS11D*) expression in the SG and gingival mucosa (Figure 4a,b) using *in-situ* hybridization (ISH) and immunofluorescent (IF) imaging (Figure 4c; Supp. Figure 4a,b). As expected, both the minor and major SG express viral entry receptors in acini and ducts (parotid gland; Supp. Figure 4c, d). Available bulk RNAseq of the major and minor SGs was used to compare *ACE2* and *TMPRSS2* expression between different SG. Minor SGs were found to express higher *ACE2* compared to parotid and submandibular SGs assessed which was confirmed using ISH (Supp. Figure 4c-e). ISH of the gingival basal and suprabasal tissues showed more expression of *ACE2* and *TMPRSS2* family members in the suprabasal cells, compared to basal populations (Figure 4d).

**Figure 4.**
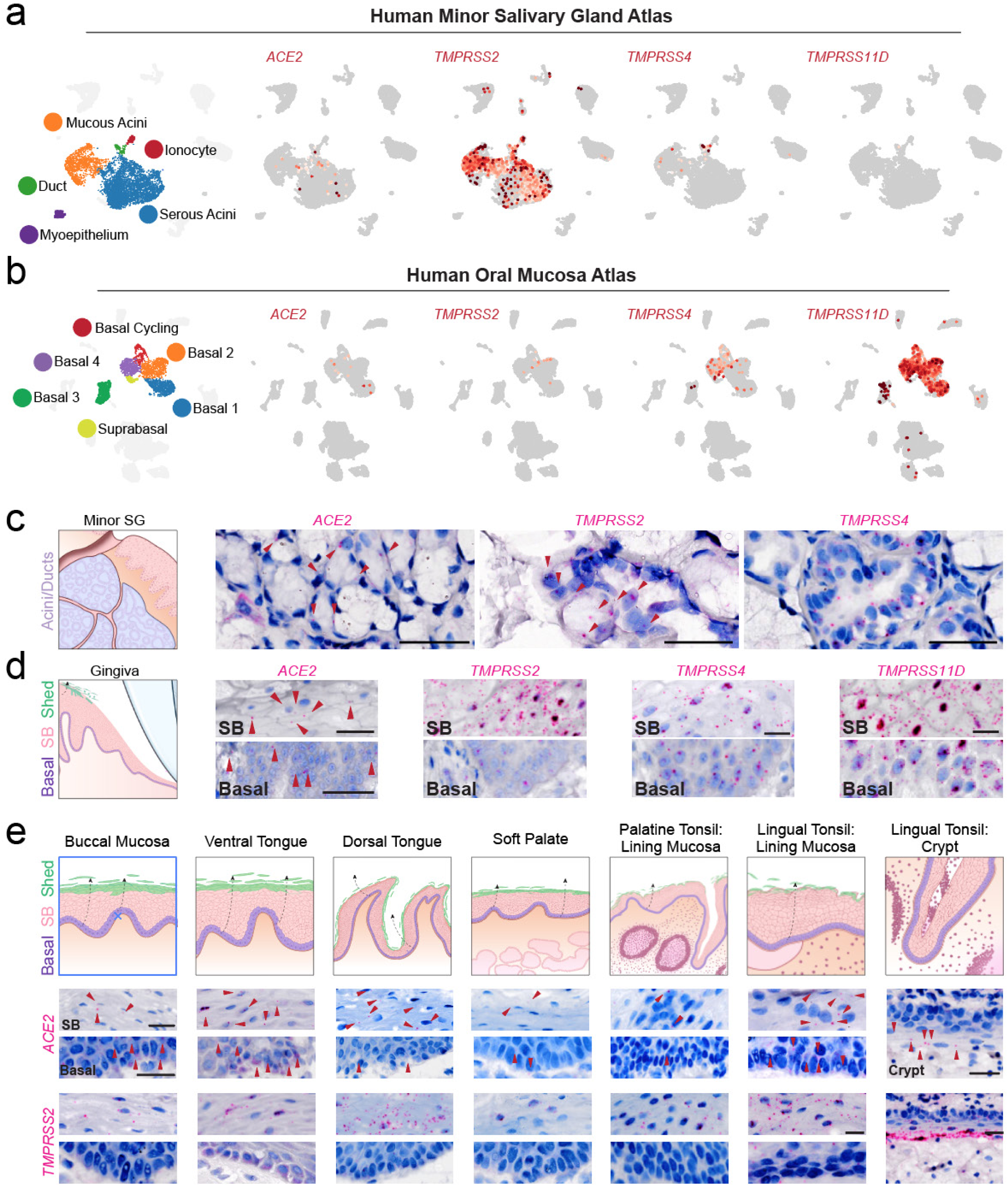
ISH across oral and oropharyngeal sites supports an oral infection axis. **(a**,**b)** UMAPs overlay of SARS-CoV-2 entry factors *ACE2* and *TMPRSS2, 4*, and *11D* in **(a)** minor saliva glands and **(b)** gingiva highlight *ACE2* expression in the saliva gland ducts and acini, and that *TMPRSS* proteases exhibits abundant expression across epithelia. Interestingly, *TMPRSS2* expression is most concentrated in the saliva glands and TMPRSS4 and TMPRSS11D are higher expressed in gingiva, with 11D displaying a defining difference between these two oral epithelial niches.**(c**,**d)** Using healthy volunteer (c) gland and (d) gingival tissue sections, mRNA expression was confirmed using RNAscope® in situ hybridization (ISH) for *ACE2, TMPRSS2, −4* as well as *-11D* in gingiva; (see *11D* in SG: Supp. Figure 4b). Due to the known shedding/sloughing of suprabasal epithelial cells (d,e; illustrations), we examined both basal and suprabasal (SB) expression, revealing enrichment of all examined entry factors in suprabasal over basal cells. **(e)** Using ISH, we map *ACE2* and *TMPRSS2* (for matched TMPRSS4 and −11D, see Supp. Figure 4) in diverse oral tissues (buccal mucosa, ventral tongue, and the dorsal tongue) as well as examining the oropharynx for the first time (soft palate, tonsils). This again supported the heterogeneity that can be found in the oral cavity—not only considering basal versus suprabasal enrichment—but also across sites. This mapping also revealed all sites are vulnerable to infection in suprabasal cells that are sloughed into saliva, a striking finding. Arrowheads in (**c-e**) indicate high expression (red), Scale bars: **(c)** 50 μm, **(d**,**e)** 25 μm.

To comprehensively explore niches without published single cell atlases, *ACE2/TMPRSS* expression was mapped across additional anatomic sites in the oral cavity, including buccal mucosa, ventral and dorsal tongue, and oropharynx, including soft palate, palatine and lingual tonsillar surface mucosa and crypts (Figure 4e; Supp. Figure 4f). For each entry factor across the examined oral and oropharyngeal sites, increased suprabasal expression was observed when compared with the basal epithelial compartment (Figure 4e). Additionally, substantial *ACE2* expression was observed in the in the tonsillar crypt epithelia. It known that the suprabasal cells of oral mucosa are continuously shed into saliva multiples times per day (49), so the expression patterning of suprabasal mucosal cells raised the possibility that sloughed cells may be infected and serve as a viral carrier to other parts of the body or to other humans (see: illustrations in Figure 4e-g). These results point to multiple sites in the oral cavity for infection and high-light oropharyngeal tissues, specifically the tonsils, as being particularly important for future study.

Droplet digital PCR (ddPCR) was used to assess SARS-CoV-2 transcripts in a cohort of COVID-19 autopsy tissues from 28 glands and 6 mucosal sites from 18 patients. SARS-CoV-2 was detected in the SG of 57% of the autopsies (16/28 glands investigated; Figure 5c; Supp. Table 1) with a trend towards higher viral loads in the minor glands than the paired parotid glands (n=8 pairs, p=0.0625, Wilcoxon 2-sided signed-rank test; Supp. Figure 5c) as predicted by bulk RNAseq. Infection by SARS-CoV-2 in the submandibular glands was also shown in 2 cases (15). SARS-CoV-2 infection of lining cells of the oral mucosa was verified in two sets of tissues from autopsy patients (P11, P23) where 5/6 of the available oral cavity mucosal sites (dorsal tongue, tonsil, uvula) exhibited detectable SARS-CoV-2 by ddPCR (Supp. Table 1), although levels of virus varied across tissue types. Thus, SARS-CoV-2 can infect SGs and mucosal sites.

**Figure 5.**
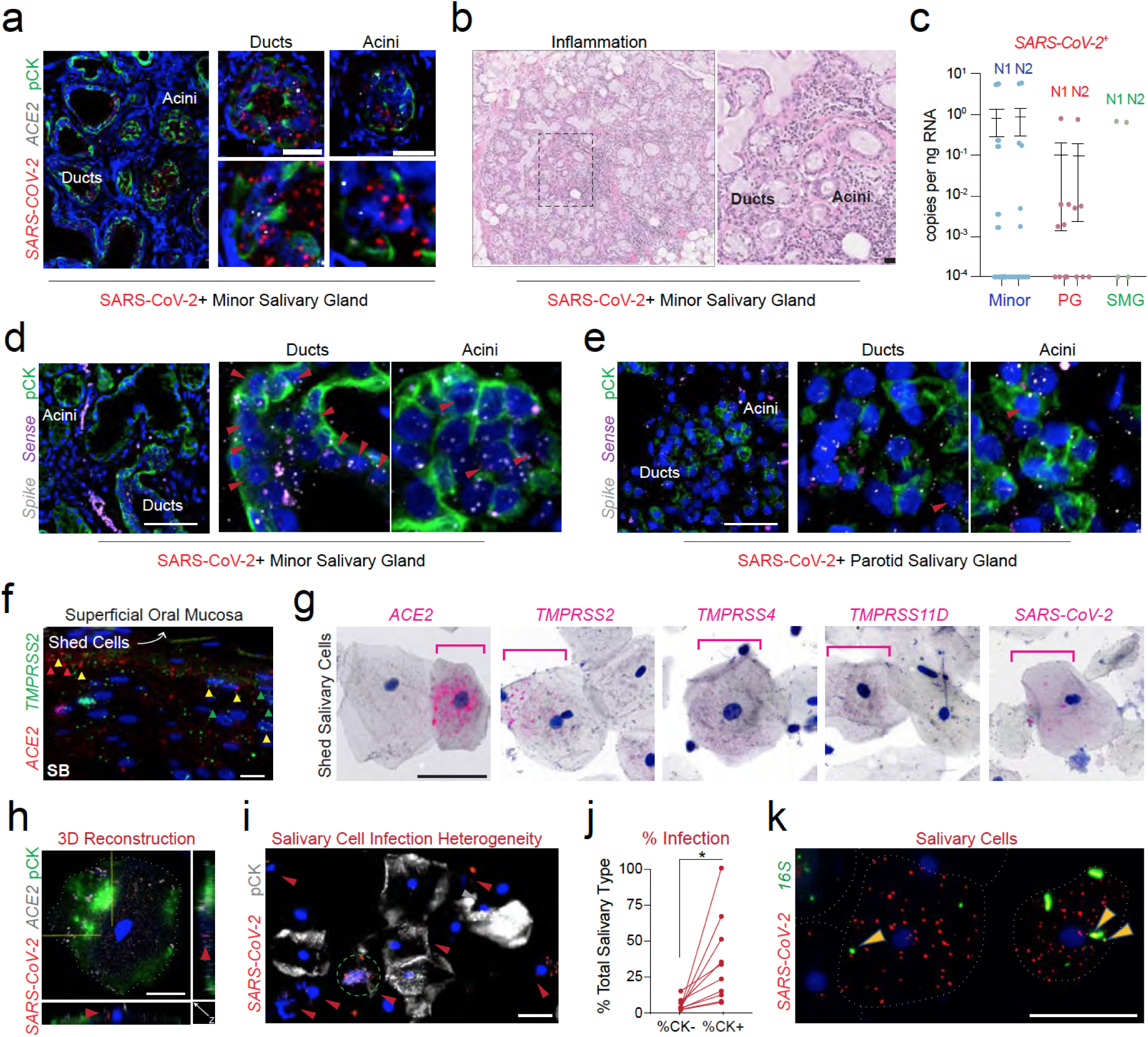
Epithelia across oral niches and in saliva are infected by SARS-CoV-2. **(a**,**b) (a)** In situ hybridization (ISH) demonstrates infection in the minor SGs (pan-cytokeratin; pCK). **(b)** H&E of COVID-19 saliva glands reveals variable histology, including variable chronic sialadenitis and some lymphocytic aggregates. **(c)** Infection was confirmed using digital droplet PCR on additional tissues recovered from 16 COVID-19 victims (12/24 in >1 SG). Minor SGs exhibited higher viral burden than parotid in paired samples (n=7). **(d**,**e)** Detection of SARS-CoV-2 infection and replication in the salivary glands using spike (V-nCoV2019-S) and sense (V-nCoV2019-orf1ab-sense) probes, respectively (arrows denote replication signal). **(f-h)** ISH of suprabasal cells confirm high concentration of ACE2;TMPRSS2 cells in cells just before sloughing/shedding. (g) Sloughed saliva cells were found to express all SARS-CoV-2 entry factors. We show that these cells can be infected by SARS-CoV-2. **(f)** This often occurs in highly expressing ACE2 cells. **(h)** Using 3D confocal microscopy, we demonstrate virus inside of these epithelial cells (see Supp. Movies 1-3). **(i**,**j)** Across saliva cells, there is infection heterogeneity of pCK cells). **(j)** Using 10 samples collected from COVID-19 outpatients (UNC OBS-C), we confirm pCK cells are the primary infected population. **(k)** SARS-CoV-2 can be found to be associated with the diverse oral microbiome on shed epithelial cells; Dotted black box in **(b)** represents zoomed-in regions; arrowheads in **(d)** replicating SARS-CoV-2 sense strands); in **(f)** indicate ACE2 (red), TMPRSS2 (green) or co-expressing (yellow) suprabasal cells; dotted white lines **(h)** highlight cell membranes; pink brackets in **(g)** represent high expressing cells; arrowheads in **(i)** indicate SARS-CoV-2+ (red) cells. Arrowheads in **(k)** indicate SARS-CoV-2 (red), universal 16S probe (green) that are co-expressing (yellow); statistical test in (i): paired Student’s t-test; * = p<0.05. Scale bars: (**a**,**b**,**d**,**e**,**h**) 25 μm, (**f**,**g**) 10μm.

To confirm tissue tropism for SARS-CoV-2 in the SG, *ACE2* and SARS-CoV-2 *Spike* ISH was completed on tissue sections from parotid (P5, P7, P19) and minor (P7, P19) SG recovered from COVID-19 autopsies. *SARS-CoV-2* was detected in *ACE2*-expressing ducts and acini (Figure 5a; Supp. Figure 5a). However, SARS-CoV-2 signals in salivary epithelia appeared regional, with some acinar units exhibiting more infection than others. The variable presence of virus in the SG raised questions whether ducts and/or acini support replication of SARS-CoV-2. Using ISH targeting the sense strand of SARS-CoV-2, minor and parotid SG (P19, Supp. Table 1) exhibit isolated clusters of acinar and ductal cells exhibiting replication. Inflammatory responses to the presence of virus in the glands ranged from mild to severe in SGs (Figure 5b; Supp. Figure 5b).

Microscopic assessment of the COVID-19 autopsy SGs revealed additional histological findings, including variable chronic sialadenitis characterized by lymphocytic aggregates or more diffuse inflammation and architectural distortion. Other samples exhibited atrophy and fibrosis, mucous inspissation, and ductal dilation. These results confirm that mucosa are sites of SARS-CoV-2 infection and establish the major and minor SG as susceptible sites for infection, replication, and distribution of SARS-CoV-2 via saliva.

### Infection of epithelial cells in saliva sets up an oral-systemic axis for viral diseases

ISH studies confirmed that the stratified squamous cells of the lining mucosa of the oral cavity and oropharynx shed into saliva expressed *ACE2* and *TMPRSS* family members required for SARS-CoV-2 infection (Figure 5d,e; Supp. Figure 5e,f). This finding raises two critical questions concerning the role(s) of the oral mucosa in the pathogenesis of COVID-19.

Firstly, because principal entry factors are expressed in the suprabasal cells of the oral mucosa that are exposed to the environment, it is possible that SARS-CoV-2 infected cells may be first infected i*n situ*, then shed into saliva, and contribute to the virus in saliva for transmission. Second, it is possible that populations of shed infected cells may provide binding sites for SARS-CoV-2 and/or act as carriers/adjuvants to promote viral stability and transmissibility.

To explore these possibilities, saliva samples collected from asymptomatic and mildly symptomatic SARS-CoV-2 positive subjects were analyzed for SARS-CoV-2 entry factor expression and found to contain both epithelial (pan-cytokeratin positive; pCK+) and immune (pCK-) populations. ISH demonstrated heterogeneity, ranging from no to high expression of *ACE2* in nucleated squamous keratinocytes (Figure 5f,g). Likewise, *TMPRSS2, −4, and −11D* were variably expressed (Figure 5g). Low levels to no *ACE2* expression were detected in the immune cells (pCK-; Supp. Fig 6a,b). Further analysis by ISH on sloughed epithelial cells in COVID-19 saliva samples demonstrated SARS-CoV-2 *Spike* expression, often in the *ACE2* expressing cells (Figure 5g; Supp. Figure 5g). To quantify this phenomenon, a 10-subject cohort (UNC) showed significantly more SARS-CoV-2 signals in pCK+ cells (34.1% pCK+ vs. 3.5% pCK-; range 5.5-100% and 0.39-13.56%, respectively), supporting the possibility of mucosal infection in SARS-CoV-2 oral pathogenesis (Figure 5g,h). While reports suggest about 50% of shed cells in saliva are epithelial (66), these data show that only ∼25% of shed cells in COVID-19 subjects were epithelial (range 4.0-39%). This reduction likely reflects a shift in the proportions of cell populations due to immune response or dropout of epithelia due to programmed cell death denoted by sparse cytokeratin expression (Figure 5g). Although speculative, this cell type may account for the rare pCK-, SARS-CoV-2-positive population. This conclusion was supported by UMAPs of salivary neutrophils, a dominant population of immune cells found in saliva, which demonstrated virtually no *ACE2* or *TMPRSS2* expression (Supp. Figure 6a; 50). A rare population of pCK-positive *ACE2*-expressing SARS-CoV-2-positive ciliated cells were found in saliva (Supp. Figure 5h) indicating that a very small fraction of saliva viral load reflects contributions from the nasal cavity. Notably, SARS-CoV-2 *Spike* independently localized both inside the sloughed squamous epithelial cells, consistent with infection, but also on the membrane surface (Figure 5i; Supp. Movies 1,2). A universal 16S probe revealed occasional colocalization of oral bacteria and SARS-CoV-2, highlighting the need to better understand the shed epithelial cell microbiome (Figure 5i).

**Figure 6.**
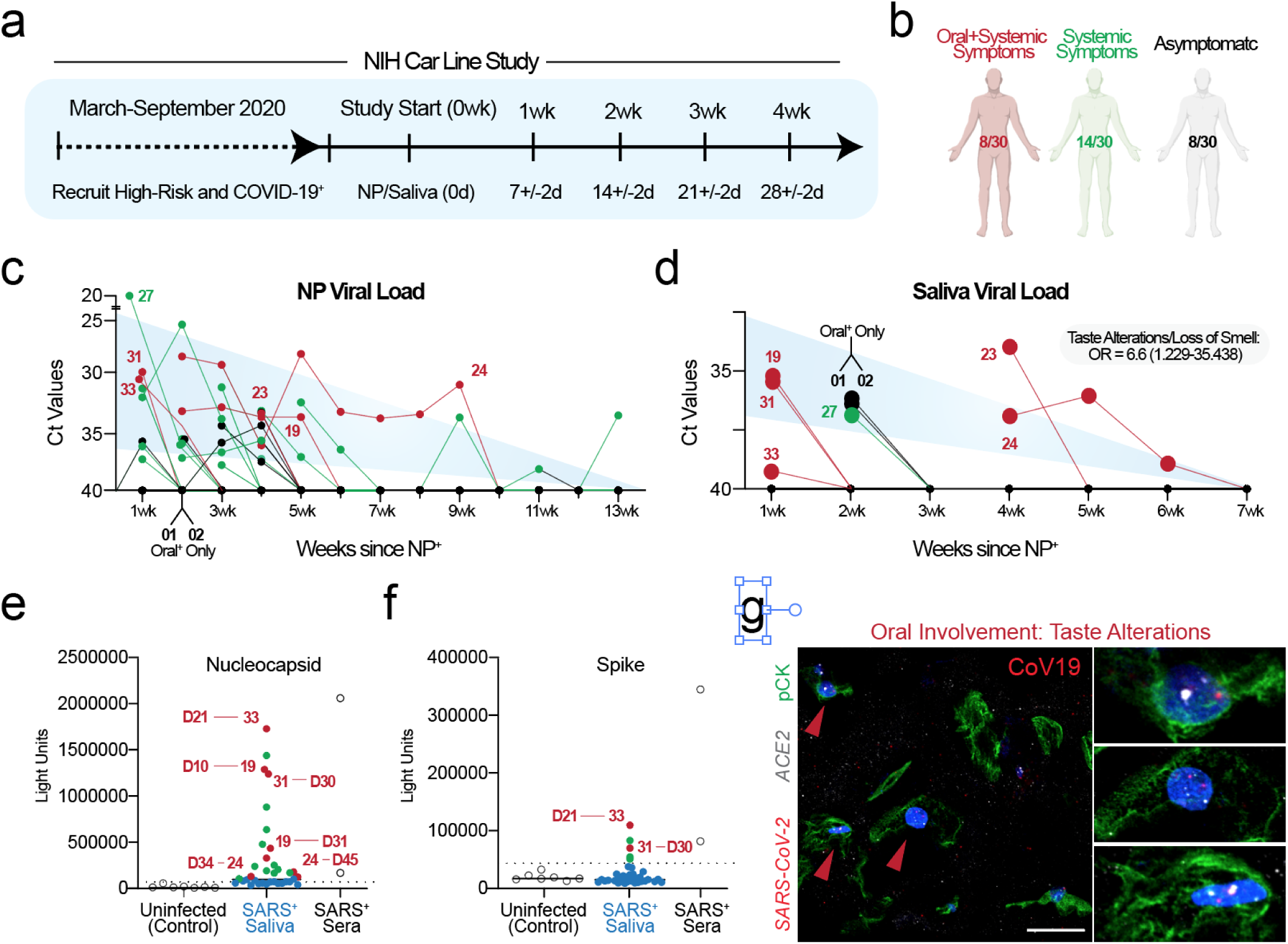
An oral infection axis is revealed by sampling high-risk and COVID-19 subjects. **(a**,**b) (a)** Schematic outlining the NIH Transmissibility and Viral Load of SARS-CoV-2 Through Oral Secretions Study (AKA - “NIH Carline Study”) design for prospective sampling of nasopharyngeal (NP) swabs and saliva over time. **(b)** Patients were grouped as having oral symptoms (with or without systemic symptoms), systemic symptoms only, or asymptomatic. **(c**,**d) (c)** Subjects demonstrated a heterogeneous disease course, with some requiring up to 13 weeks to clear detected virus. **(d)** The majority of asymptomatic subjects only displayed NP swab positivity, including two patients who developed symptoms during the prospective sampling (CoV12 and CoV13; see Supp. Figure 6). However, while CoV01 and Cov02 were enrolled in the study due to a positive NP swab, and for the first time, we show the possibility of oral infection without maintaining NP infection. **(e**,**f) (e)** Evaluation of a pilot set of saliva samples for nucleocapsid and spike antibodies by Luciferase Immunoprecipitation System (LIPS) assay shows saliva from a cohort of COVID-19 patients (N = 36) and healthy control individuals (N = 7) contains antibodies to both viral antigens. Seropositive nucleocapsid antibodies occurred in 50% (18/36 subjects) of the COVID-19 cases and showed a range of antibody levels. **(f)** In contrast, seropositive spike antibodies were less common with a frequency of 14% (5/36; Figure 6g). **(g)** In patients with the highest salivary viral load and reported taste alterations (dysgeusia in CoV19), we observe significant salivary epithelial (pan-cytokeratin positive) cells; see also Supp. Figure 6e,f). Scale bars: (**a**,**e**) 25μm.

In addition to virus dispersion from droplets and aerosols of saliva, these data emphasize the potential role for shed epithelial cells in saliva to deliver virus to other mucosal sites of the body, such as the gastrointestinal tract (51, 52) and lungs (53). Using ISH to detect SARS-CoV-2 RNA suggestive of viral replication (*V-nCoV2019-S* sense probe), both minor and major SG demonstrated clusters cells containing replication (P19, Figure 5d,e). Thus, these results point to two oral innate immune responses to viral challenge. While broadly susceptible to infection, the mucosa may successfully shed infected cells and downregulate transcriptional machinery. Alternatively, while less accessible and protected by outward salivary flow, once infected the SG may result in a more established and symptomatic infection.

### SARS-CoV-2 in saliva correlates with symptoms and supports an oral axis for COVID-19 pathogenesis

To investigate the possibility of SARS-CoV-2 in asymptomatic transmission by saliva, subjects that were COVID-19 positive who were either asymptomatic or mildly symptomatic, or asymptomatic subjects at a high risk of infection were studied. Two groups of subjects were enrolled in the study (N=36): subjects with a positive NP swab (n=28); or subjects with high-risk contact (co-habitation, occupational exposure, n=8), two of which tested positive at the first study visit (Figure 6a,b). Over a 5-week prospective interval, this trial studied the presence of SARS-CoV-2 in saliva and nasopharyngeal (NP) swabs and the presence of antibodies against viral antigens in saliva. Based on the time from their first positive NP swab test, these data confirmed what others have recently reported54, i.e., that salivary and NP swabs generally correlated for SARS-CoV-2 positivity over time (Figure 6c,d). However, these data also highlighted novel points in this outpatient cohort. First, some subjects cleared the virus over very long periods of time (>2 months since first test to negative NP and saliva tests). Second, not all truly asymptomatic individuals rapidly clear virus from their secretions. Of the 36 subjects enrolled in this study to date, 25% reported no symptoms at all (8/30) with a range of viral of clearance from 0.5 weeks to 3.5 weeks (average 18 days after first positive NP swab). This rate is only one week less than in subjects with symptoms, suggesting that some asymptomatic subjects may carry virus in their nasopharynx and saliva over extended periods of time. Out of 8 entirely asymptomatic participants, 5 were positive only in their NP swabs including two who developed positive viral loads in the middle of the monitoring period (Supp. Figure 6c). Two asymptomatic participants, CoV01 and CoV02, displayed only positive salivary viral signals when matched to NP viral loads 14 days after the positive NP test (Figure 6d). This data highlights that it is possible to clear virus from the NP while displaying persistent virus in saliva suggesting sustained shedding of virus from SG or from SARS-CoV-2 infected epithelial cells.

The association of oral infection and COVID-19 symptom was also examined. In symptomatic subjects, the presence of SARS-CoV-2 RNA in saliva was positively associated with reported loss of taste and smell (OR: 6.6 [1.229 – 35.438]; p<0.05; Figure 6d; Supp. Table 2) indicating viral infection of cells of the taste papillae. Moreover, these data are supported by the saliva cell infection experiments (Supp. Figure 6b) showing two patients with high viral load in saliva and reported taste changes displayed significant epithelial (pCK+) cell infection in *ACE2* expressing cells (Figure 6h, Supp. Figure 6e,f). Reports of “body ache/muscle pain” was inversely correlated to saliva viral burden (OR: 0.090 [0.009 − 0.880]; p<0.05; Supp. Table 2) indicating differential patterns of infection and immune response.

Public health measures, such as universal mask use and social distancing, are intended to reduce droplet transmission. However, few studies have attempted to directly measure the change in saliva droplets ejection from COVID-19 subjects by wearing a mask. The effectiveness of standard mask wearing to reduce droplet spread in these subjects was tested and demonstrated decrease in the detection of expelled droplets >10-fold (p<0.005), including some asymptomatic subjects with positive NP or saliva viral load; although the assay did not have the sensitivity to detect SARS-CoV-2 RNA (Supp. Figure 6d).

Saliva is a complex fluid that functions to protect the oral cavity tissues through a variety of mechanisms, including secreted salivary proteins and antibodies (IgG, IgM, and IgA) which potentially restrict viral infections (55). However, little is known about the local antibody responses at the site of infection in the oral cavity for SARS-CoV-2 and only very recently has the oral cavity been tested for antibodies to SARS-CoV-2 antigens (56). Utilizing the Luciferase Immunoprecipitation System (LIPS) assay, antibodies were detected to the viral antigens nucleocapsid and spike in 50% (18/36) and 14% (5/36), respectively, in early recovered COVID-19 subjects (Figure 6f,g). Surprisingly, several of the subjects with both nucleocapsid and spike proteins were those individuals with protracted positivity in NP and saliva, moderate symptoms, and indica-tors of oral infection (ex: CoV19 in Figure 6h; CoV23 in Supp. Figure. 6e). The presence of antibodies in saliva may indicate the presence of local infection in oral cavity tissues. It is now known that anti-SARS-CoV-2 antibodies exhibit a temporal relationship with nucleocapsid antibodies developing prior to the detection of spike antibodies (57). Due to the short-term window of this study, participants were requested to return at 6-months and 1-year for antibody surveillance in serum and saliva.

## DISCUSSION

Collaborative efforts led by the Human Cell Atlas (HCA) utilized single cell RNA sequencing (scRNAseq) datasets from across the body to examine cell-specific SARS-CoV-2 tropism, leading to the COVID-19 Cell Atlas (https://www.covid19cellatlas.org/). These and other atlases point to the permissiveness of the barrier epithelium to SARS-CoV-2 infection (58, 59). While viewed as an important tool, oral scRNAseq datasets were not included in these efforts. To address this gap in knowledge, the studies herein are the first integrated adult human oral scRNAseq atlas and these studies suggest a range of oral infection susceptibilities that may medi-ate the oral axis in COVID-19 pathogenesis and transmission. Although principally focused on COVID-19, these data emphasize the unique ana-tomic and cellular diversity found in the oral cavity and underscore future opportunities to complete a comprehensive oral and oropharyngeal atlas (Figures 1b, 4; 26, 28).

The COVID-19 integrated oral atlas focused our investigations on the spe-cialized oral epithelial populations, including first the “secretory” epithelia of the major and minor SGs (Figure 2; Supp. Figure 1). The SGs exhibit enriched, albeit low, expression of principal entry factors in the epithelia and co-expression of *ACE2* and *TMPRSS2* was found in 0.54%, 0.23%, and 0.71% of mucous, ductal, and myoepithelial cells, respectively. These percentages are slightly lower than reported for cells in the respiratory tract but may reflect limitations in the detection of low-expression genes by scRNAseq. Co-expression of these markers was more frequent using ISH and emphasized that SARS-CoV-2 is predicted to have tropism to the acini and ducts. While there has been a historical focus on major SG viral infections, broad expression of viral entry factors and significant infection of minor SG epithelia was confirmed. Minor SGs are widely distributed among the tongue, palate, and mucosae (60), sites now demonstrated to be hotspots for SARS-CoV-2 infection. The novel finding of SG and tongue infection helps explain acute onset and post-recovery COVID-19 symptoms, such as dry mouth and taste alterations (14); however, infection and productive replication in even a majority of these glands may produce no symptoms, possibly explaining the phenomenon of “silent spreaders” in COVID-19.

One limitation of our scRNAseq approach was the cellular representation of the mucosal atlas which enriched for immune cells over suprabasal squamous epithelial cells (Figure 2; Supp. Figure 1). When compared to the SG, these results suggested that SARS-CoV-2 would have relatively little tropism for the mucosa based on the low expression and abundance of cells expressing *ACE2* and *TMPRRS2*. However, integrated pan-oral mouse single cell data revealed a more robust description of the oral cavity due to increased numbers of suprabasal cells from the tongue (47) and buccal mucosa (46), which exhibited higher levels of *Ace2* and *Tmprss2* expression compared to basal epithelial cells. Examination of human gingiva and other oral niches and found that the Basal 1 population exhibited co-expression of *ACE2* and *TMPRSS2* in 0.3% of cells, similar to our ductal cell percentage. Basal cells eventually delaminate from the suprabasal layer and shed into saliva (Figure 4; 28). Surprisingly, as delaminating basal cells differentiate into suprabasal cells, their expression of *ACE2* and *TMPRSS* family members increased as they were poised to shed into the oral cavity (Figure 4). This finding suggests the possibility that in some cases COVID-19 occurs without symptoms since stratified squamous cells are not linked to specific symptoms and are continuously shed into saliva (Figure 5,6; 49. These infected, shed oral epithelial cells can elicit a ro-bust immune response in the mucosa and gut leading to local production and secretion of antibodies via the oral mucosae and SGs (63). This axis explains the relationship between oral infection and the development of robust antibodies detected in saliva in the clinical dataset (i.e., CoV19, CoV33, Figure 6c-h).

Two independent and complementary datasets from outpatients revealed the presence of SARS-CoV-2 infected squamous epithelial cells in saliva, providing a potential cellular mechanism for disease spread and transmission. The discovery of an oral source of infection and replication in the SG as well as the natural conduit for viral spread via saliva establishes the possibility of two infection axes in COVID-19 and highlights how simultaneous testing of oral and nasopharyngeal sites might be required to fully understand SARS-CoV-2 spread (Figure 1a). However, these results raise new questions about COVID-19 pathogenesis including: 1) whether this is primarily a “nasal-first” infection that spreads via mucus transport to the oral cavity or 2) the possibility of an “oral-first” infection via fomite or droplet inoculation, and 3) whether the pattern of infection impacts disease severity and host immunological responses. To test whether oral transmission can precede nasal and/or occur in the absence of nasal infection, it will be necessary to design studies that include daily surveillance using NP and salivary tests in an at-risk cohort.

Considering now-documented oral SARS-CoV-2 infection and the ease of saliva for transmission, it remains critical to further our understanding of the dominant modes of viral spread across the spectrum of asymptomatic, pre-symptomatic, and symptomatic individuals. Asymptomatic transmission of SARS-CoV-2 is the “Achilles’ Heel” of this pandemic (64), and asymptomatic or pre-symptomatic spread has been estimated to cause to 2.2-45% of COVID-19 cases. COVID-19 infections are now widely accepted to involve airborne droplet transmission based on guidance from the WHO and CDC. However, the presence of SARS-CoV-2 infectious particles in the SGs, the oral mucosa, and in saliva raises the possibility that the oral cavity actively participates in SARS-CoV-2 infection and transmission (Figure 1a). Finally, these data provide strong evidence in support of universal public health measures, including mask wearing, social distancing, and hand washing, to limit exposure to potentially infectious droplets, aerosols, and fomites generated from the oral cavity (Supp. Figure 6d).

## MATERIALS AND METHODS

### HUMAN GINGIVAL SINGLE CELL RNA SEQUENCING

Clinical Protocol: Five subjects were enrolled in this study conducted at the University of North Carolina Adams School of Dentistry (Chapel Hill, North Carolina), and 5 total healthy subjects (2 females and 3 males; aged 20 to 30 years) completed the experimental gingivitis protocol in October 2019. Each subject was determined to be systemically healthy individuals with mild gingivitis. Subjects received initial prophylaxis (Day 0) with a 3-week stent-induced, biofilm overgrowth, experimental gingivitis induction phase over the upper left maxillary premolar and first and second molars only (Clinicaltrials.gov ID: NCT04105569; UNC IRB: 19-0183). Full-thickness gingival biopsies (0.3μm3) were collected on the same day from each subject at the 3-week time point from the upper left maxillary lingual interproximal papilla (universal number #14-15): Biopsy samples were split for tissue dissociation and tissue fixation.

Cold Protease Dissociation/FACS: Split tissues were placed immediately in ice cold, calcium-free AND magnesium-free 1x PBS for overnight shaking and incubation with 1:1000 ROCK Inhibitor (Sigma-Aldrich; Y27632). Cold protease dissociation cocktail was made using cold 1x PBS, 2x DNASE (2.5mg/mL Sigma-Aldrich; 9003-98-9), and 1:1000 ROCK Inhibitor. Protease (Sigma-Aldrich; P5380) was then added to a concentration of 100mg/mL. Sterilized dissecting forceps (Roboz) were used to mechanically separate gingival into several pieces. Enzymatic cocktail was incubated with gingival tissues for 35 minutes but progress was checked every 10 minutes with 10μL. Dissociation was completed when > 75% of the solution was single cells, then gingival single cells were pelleted at 1800 rcf and washed twice with an appreciable volume of a cocktail of cold culture medium, 1:1000 Y-27632, and 10% FBS. Following dissociation, viability was checked using trypan blue (viability was confirmed >80%) and RBC lysis was achieved using a 1x RBC buffer (Biolegend 420301). A FACS buffer (mixture of PBS, 1% FBS, and 1:1000 ROCK Inhibitor) was used throughout the resuspension and cell sorting process. Single cells were enriched using a 35mm Falcon cell strainer (Thermo Fisher), Before FACS, the cell pellet was resuspended in the FACS buffer and stored on ice. To isolate living cells by FACS, Rat anti-CD49f-Alexa Fluor 647 (a6-In-tegrin, BioLegend clone GoH3) was added to cells and incubated for 60 min on ice. Sytox (Thermo Fisher, 1:1000) was added 5 min before FACS to evaluate live/dead cells using a Sony SH800S Cell Sorter, with the following controls: anti-CD49f:Alexa Fluor 647 negative, and Sytox Dead Cell Stain negative cell suspensions. Cells were sorted ∼6 counts/sec, and live cells were sorted into tubes directly for 10x capture.

10x Capture, Library Prep, Sequencing: Cells were processed using the 10x Genomics Chromium Controller and the Chromium Single Cell 3’ GEM, Library & Gel Bead Kit v3 (PN-1000075) following the manufacturer’s user guide (https://tinyurl.com/y28bwe67). Briefly, aliquots of the sorted cells were stained with acridine orange and propidium iodide and assessed for viability and concentration using the LUNA-FL Dual Fluorescence Cell Counter (Logos Biosystems). Approximately 3,200 cells per sample were loaded onto the Chromium Chip B with a target recovery of 2,000 cells per sample for library preparation. Single cells, reverse transcription reagents, and gel beads coated with barcoded oligos were encapsulated together in an oil droplet to produce gel beads in emulsion (GEMs). Reverse transcription was performed using a C1000 thermal cycler (Bio-Rad) to generate cDNA libraries tagged with a cell barcode and unique molecular index (UMI). GEMs were then broken, and the cDNA libraries were purified using Dynabeads MyOne SILANE (Invitrogen) prior to 12 amplification cycles. Amplified libraries were purified with SPRIselect magnetic beads (Beckman Coulter) and quantified using an Agilent Bioanalyzer High Sensitivity DNA chip (Agilent Technologies). Fragmentation, end repair, A-tailing, and double-sided size selection using SPRIselect beads were then performed. Illumina-compatible adapters were ligated onto the size-selected cDNA fragments. Adapter-ligated cDNA was then purified using SPRIselect beads. Uniquely identifiable indexes were added during 12 amplification cycles. The finalized sequencing libraries were then purified using SPRIselect beads, visualized using the Bioanalyzer High Sensitivity DNA chip, quantified with the KAPA SYBR® FAST Universal qPCR Kit for Illumina (Roche) and StepOnePlus Real-Time PCR System (Applied Biosystems), normalized to 4 nM, and pooled. Pooled libraries were sequenced on a NextSeq 500 machine (Illumina). Libraries were denatured and diluted following standard Illumina protocol, spiked with 1% PhiX sequencing control (Illumina), loaded onto the flow cell at 1.8 pM, and sequenced in paired-end format (Read 1: 28 bp, Read 2: 91 bp) to a total depth of 523M read pairs passing quality filters. The 10x Genomics Cell Ranger analytical pipeline v3.1.0 and GRCh38 v3.0.0 reference package were used to analyze the data following default settings.

### HUMAN MINOR SALIVARY GLAND SINGLE CELL RNA SEQUENCING

**T**issue Dissociation: Minor SGs were collected from healthy volunteers who provided informed consent on NIH protocols 15-D-0051 or 94-D-0094. De-identified submandibular and parotid s were obtained fresh from the Human Cooperative Tissue Network and portioned into RNAlater for RNA sequencing or were formalin-fixed in 10% neutral buffered formalin overnight before being transferred to 70% PBS-buffered ethanol (Hewitt reference), then paraffin-embedded for immunohistochemistry and in situ hybridization studies. Five healthy subjects provided minor s biopsies for single cell RNA sequencing. Tissues were biopsied following standard methods and immediately placed in ice cold RPMI. Tissues were sharply dissected into 1-2 mm pieces then dissociated using the Miltenyi Multi-tissue Dissociation Kit A using the Multi_A01 in C-type tubes at 37C in an octoMacs tissue disruptor using heated sleeves. Crude single cell suspensions were serially filtered through 70 and 30 uM filters and rinsed with 1X-Hank’s Buffered Salt Solution. Cells were centrifuged at 300g for 10 minutes at 4C and washed once with 1X Hank’s Buffered Salt Solution. Cell counting and viability was determined using a trypan blue exclusion assay. Suspensions with greater than 35% viability were used for subsequent sequencing. Additional glands from these patients were submitted for histopathological assessment including focus scoring (DEK).

Single-cell capture, library preparation and sequencing: Single cell suspensions targeting approximately 5,000 cells were prepared as described above and loaded onto a 10X Genomics Chromium Next GEM Chip B following manufacturer’s recommendations. After cell capture, single-cell library preparation was performed following the instructions for the 10× Chromium Next GEM Single Cell 3’ kit v3 (10x Genomics). The libraries were pooled and sequenced on four lanes of a NextSeq500 sequencer (Illumina), adopting the read configuration indicated by the manufacturer. Single-cell RNA-seq data processing and quality control: Read processing was performed using the 10x Genomics workflow. Briefly, the Cell Ranger v3.0.1 Single-Cell Software Suite was used for demultiplexing, barcode assignment, and unique molecular identifier (UMI) quantification http://software.10xgenomics.com/single-cell/overview/welcome). Sequencing reads were aligned to the hg38 reference genome (Genome Reference Consortium Human Build 38) using a pre-built annotation package obtained from the 10X Genomics website (https://support.10xgenomics.com/single-cellgeneexpression/software/pipelines/latest/advanced/references). Samples were demultiplexed using the ‘cell ranger mkfastq’ function and gene counts matrices were generated using the ‘cellranger count’ function. The single cell data was analyzed in R (v3.5.0) using Seurat (v3.1.2). Filtering was performed using the standard quality control steps provided on the Satija Lab website (satijalab.org). Cells containing more than 200 and less than 2,500 unique features were retained. From this set, cells with greater than fifteen percent of read counts attributed to mitochondrial DNA were filtered out. We adjusted this value from five to fifteen percent to increase the yield from each sample and did not observe substantive changes in our results after adjustment. Data were normalized using the “NormalizeData” command (scale factor = 10,000).

Annotation of clusters: SG cells from n=5 non-Sjögren’s syndrome and otherwise healthy subjects were integrated into a single Seurat object containing 12 clusters at resolution=0.1. The clusters were manually annotated with marker genes identified by Seurat’s “FindAllMarkers” function, using the receiver operator characteristic (ROC) test. The “FeaturePlot” function was used to identify subpopulations within identified clusters and confirm the expression of known marker genes within clusters.

### INTEGRATED SINGLE CELL ANALYSES

Clustering and Annotation: Oral atlases were retrieved from published and unpublished datasets (human salivary glands: Warner et al.; human gingiva/palatal mucosa: Byrd et al.). Raw expression matrices were filtered, normalized and log transformed for further processing using standard SCANPY (v.1.4.3) procedure (68). For the published mouse datasets, we considered original publication annotations when re-processing and annotating cell types. For the two unpublished human datasets, BBKNN was used for batch correction across samples and multiple clustering rounds of different resolutions were performed to resolve subpopulations and then manual annotations were made considering published human and mouse references (69). For integration across datasets, each individual dataset was first randomly down-sampled to having <=500 cells per cell type, then pooled raw expression values were re-processed following standard SCANPY procedure while using Harmony for correcting batch effects across datasets and samples (70). Again, multiple clustering rounds of different resolutions were performed, and manual annotations were made. Illustration of the results were generated using python code around SCANPY.

### IHC AND ISH FOR HUMAN ORAL TISSUES AND SALIVA

#### Oral and Oropharyngeal Tissue Acquisition

Participant tissues included remnant oral specimens previously collected through the University of North Carolina Adams School of Dentistry Oral Pathology Biobank (UNC IRB: 20-1501) or the Center for Oral and Systemic Diseases (COSD) Biorepository (UNC IRB: 15-1814). Lab and project personnel were double-blinded and did not have access to PHI or any links to any PHI. Project members did not have interpersonal contact with patients that provided specimens. All de-identified tissues were previously fixed in 10% formalin and embedded in paraffin blocks.

#### Saliva cytospin

Saliva samples were collected from COVID-19 patients, and fixed with 4% paraformaldehyde for 30 min. Then the samples were centrifuged, and the cell pellet was dissolved into 5mM EDTA in PBS. The portion of the samples were loaded into disposable sample chambers (Double cytofunnel, Thermo Scientific, Waltham, MA, USA) with slides and applied into a cytospin machine (Shandon cytospin 4, Thermo Scientific, Waltham, MA, USA) following the manufacturer’s instructions and centrifuged at 1000 rpm for 5 min. The cytospin slides were stored into 70% ethanol under 4C until IHC or RNA ISH.

#### Saliva cell blocks

Saliva samples were diluted in 10 volumes of 4% paraformaldehyde-1XPBS. Cells were allowed to fix for 24-hours at room temperature. After 5 minutes of centrifugation at 1000xG RPM, cell pellets were transferred to 70% ethanol. Cell pellets were then ensnared by adding thrombin and fibrin to the cell pellet which could be subsequently embedded for tissue sectioning.

#### ISH/IHC

FFPE tissue sections were baked overnight followed by deparaffinization with xylene for 5 minutes twice and dehydration with 100 % ethanol for 1 minutes twice. Then, the sections were incubated with hydrogen peroxide for 10 minutes and antigen retrieval was performed with boiled water for 15 minutes and with protease at 40C for 15 minutes. Probes were hybridized at 40C for 2 hours in an oven followed by signal amplification and washing steps. The hybridized signals were visualized by Fast Red followed by counterstaining with hematoxylin. The probe targeting human housekeeping gene Ubiquitin C (UBC) was used as a positive control to test for RNA quality in the tissue sections. A bacterial gene, Bacillus subtilis dihydrodipicolinate reductase (DapB) was used as a negative control target. The images were acquired using an Olympus VS200 slide scanner light microscope with a 60x 1.42 N.A. objective. All RNAscope® in situ hybridizations were performed following the manufacturer’s recommended conditions with the following exceptions: for in situ hybridization pretreatment conditions were modified as follows: protease plus digestion at 40C for 15 min. Antigen retrieval in RNAscope® target retrieval in slide steamer control temperature at 99C for 15 min.

#### UNC

Positive control probe: 320861; Negative control probe: 320871; ACE2 antibody: Mouse Monoclonal (protein tech #66699-1-IG), (ACE2 diluted 1/2000, PBS 1% BSA, 1% donkey serum PBS); SARS-CoV-2 probe: ACD, 848561; ACE2 probe: #848151-C3; TMPRSS2 probe: 470341-C2; CSTL probe: 858611-C3; Secondary antibodies: Al secondary antibodies were in donkey (D-anti Goat, D-anti Rabbit, diluted 1/300 in 1/300 PBS 1% BSA, 1% donkey serum, PBS). For the detection of anti-ACE2 we used TSA detection kit from Biotium #33000); AQP5 antibody: Goat antibody, Santa Cruz, C-19, sc9891 (dilution 1/300 PBS 1% BSA, 1% donkey serum, PBS); ACTA2 antibody: Goat, Novus Biological, NB300-978 (Dilution 1/200 PBS 1% BSA, 1% donkey serum, PBS); pCTK antibody: Rabbit polyclonal (Abcam, ab234297); SARS2 antibody: Rabbit polyclonal, Invitrogen, Cat #PA1-41098 (1:500 dilution); Cytokeratin antibody: Mouse monoclonal, Dako Cat#M3515; Mounting Media. The slides were mounted in Prolong Gold anti-fade (with DAPI for the case if the IF and without DAP for the ISH).

#### NIH

ACE2 antibody: Mouse Monoclonal (protein tech #66699-1-IG), (ACE2 diluted 1/2000, PBS 1% BSA, 1% donkey serum PBS); validated for IF, WB, FC, and on human tissues (colon). AQP5 antibody: Goat antibody, Santa Cruz, C-19, sc9891 (dilution 1/300 PBS 1% BSA, 1% donkey serum, PBS); validated by WB, IHC on human tissues (human salivary gland with primarily apical membrane expression on acini). ACTA2 antibody: Goat, Novus Biological, NB300-978 (Dilution 1/200 PBS 1% BSA, 1% donkey serum, PBS); Validated using WB of mouse and human duodenum, and frozen section immunofluorescence of mouse heart. Expected staining is demonstrated on the terminal acini of the human salivary glands. pCTK antibody: Rabbit polyclonal (Abcam, ab234297), demonstrated reactivity with human epidermis.

V-nCoV2019-S (848561) is an antisense probe specific to the Spike protein sequence of the viral RNA so will give information about infection and viral load in tissue, i.e. the presence of virus.V-nCoV2019-orf1ab-sense (859151) (40 probe pairs) detect replicating virus. Therefore they can be used to detect viral replication in tissue. 848561 Was used in the original set of ISH (Figure 4); and in last set was mixed with 859151 (Figure 5) to detect both the virus and replication at the same time.

### CLINICAL STUDIES

#### UNC Epidemiological and Immunological Aspects of COVID-19 Saliva

A subset of 10 samples from COVID-19 outpatients were collected from an IRB approved study (PI: Bowman; UNC IRB Number: 20-0792). The purpose of this study is to describe the epidemiology, clinical features, and immunological response to SARS-CoV-2 infection and develop new diagnostic tests focused on saliva. Subjects were individuals >18 years old who are tested for COVID-19 (NP swab) at UNC Healthcare and contacted as outpatients for sample collection. The subjects in this study were classified as having “mild” COVID-19. Outpatients were enrolled during diagnosis at the UNC-CH Respiratory Diagnostic Center (RDC) and recruited no more three days following a positive clinical test. All participants were asked to come to the RDC for saliva collection, which occurred by the participant actively drooling into a 5mL collection tube. For all samples, specimens were evaluated for volume, total cell count, and cell differential before proceeding to fixation and ISH/IHC analyses.

#### NIH Transmissibility and Viral Load of SARS-CoV-2 in Oral Secretions

Subjects providing informed consent to 20-D-0094 (clinicaltrials.gov NCT04348240) were seen between the hours of 8:30am and 11:00 am the NIH COVID-19 Testing Center. Before arriving at the testing facility, subjects were asked to refrain eating 90 minutes before testing. First, subjects were asked to speak by reading Samantha Smith’s “Ambassador of Peace” address (∼2-minute duration, 5th Grade Reading Level) directly into an emesis bag shortened to a depth of 10 cm with a 1/4 section of 3”x3” Pro-Gauze® Pads (Curad®) dampened with 2 mL of 1X-PBS. Gauze pads were recovered for saliva and viral burden analysis as described above. Subjects then repeated the speaking exercise with a disposable procedure mask on day 1; day 8; and/or 15 (or day 21, or day 28, when applicable. Whole unstimulated saliva was collected in sterile 50 mL conical vials by asking patients to refrain from swallowing, allowing the pooling of saliva under the tongue and in the vestibules of the mouth then pushing the saliva into the open conical vial for 2-3 minutes. A synthetic Copan swab (NP swab) was used to transfer a standardized amount of saliva from the 50 mL tube to a new 15 mL conical tube containing 3 mL of viral transport media for CDC SARS-CoV-2 testing. Swabs of the nasopharynx (NP), were obtained and placed into 15 mL conical tubes containing 3 mL of viral transport media for CDC SARS-CoV-2 testing. Remaining samples were immediately placed on ice and transported to the NIH Department of Laboratory Medicine for testing and cryostorage. Symptom questionnaires were collected at consent, and again every 2-3 days over the course of the study.

#### CDC qRT-PCR test for testing the NP swabs and Saliva

In response to the SARS-CoV-2 pandemic, the Centers for Disease Control and Prevention (CDC) 2019 novel Coronavirus (2019-nCoV) Real-Time PCR Diagnostic Panel has received U.S. Food and Drug Administration (FDA) approval and has been adopted by the National Institutes of Health (NIH) Clinical Center, herein referred to as the SARS-CoV-2 RT-PCR assay (71). The SARS-CoV-2 RT-PCR assay is used to screen NP and OP swab specimens in VTM and BAL for SARS-CoV-2 from patients with respiratory symptoms. The SARS-CoV-2 RT-PCR assay implemented at NIH Clinical Center utilizes the easyMAG automated nucleic acid extractor (bioMérieux, Marcy l’Etoile, France). A Taqman assay using two primer/probe sets is used to detect two distinct regions of the N gene (nucleocapsid protein, referred to as N1 and N2) and an internal control to detect the human RNase P (RP1) gene present in all specimen collections. This assay was performed on the ABI 7500 Fast Real-Time PCR System (Thermo Fisher Scientific, Waltham, MA).

### PROCUREMENT AND TESTING OF AUTOPSY TISSUE SPECIMENS FOR SARS-COV-2

#### Autopsy Tissues

Consent for autopsies of COVID-19 victims was coordinated by the NIH COVID-19 Autopsy Consortium and obtained from family members. Autopsies were performed in the NCI Laboratory of Pathology and tissues were recovered in accordance with family wishes for histopathological and other downstream analyses.

#### Viral Quantification of Autopsy Tissues

Total RNA was extracted from RNAlater (Invitrogen, Hampton, NH, USA) preserved tissues collected at autopsy using the RNeasy Fibrous Tissue Mini Kit (Qiagen, Germantown, MD, USA) according to the manufacturer’s protocol. Prior to extraction, tissues were mechanically homogenized on the GentleMACS Octo Dissociator with Heaters (Miltenyi Biotec Inc, Auburn, CA, USA) using the gentleMACS M tubes with the RNA_02 program. The NanoDrop ND-1000 Spectrophotometer (Thermo Scientific, Waltham, MA, USA) was used to quantify RNA concentrations. The QX200 AutoDG Droplet Digital PCR System (Bio-Rad, Hercules, CA, USA) was used to detect and quantify SARS-CoV-2 RNA using the SARS-CoV-2 Droplet Digital PCR Kit (Bio-Rad), which contains a triplex assay of primers/probes aligned to the CDC markers for SARS-CoV-2 N1 and N2 genes and human RPP30 gene. 96 well plates were prepared with technical replicates of up to 550 ng RNA per well using the aforementioned kit according to manufacturer’s instructions. The QX200 Automated Droplet Generator (Bio-Rad, Hercules, CA, USA) provided microdroplet generation and plates were sealed with the PX1 PCR Plate Sealer (Bio-Rad, Hercules, CA, USA) before proceeding with RT-PCR on the C1000 Touch Thermal Cycler (Bio-Rad, Hercules, CA, USA) according to the manufacturer’s instructions. Plates were read on the QX200 Droplet Reader (Bio-Rad, Hercules, CA, USA) and analyzed using the freely available QuantaSoft Analysis Pro Software (Bio-Rad) to quantify copies of N1, N2, and RP genes per well, which was then normalized to RNA concentration input. For samples to be considered positive for SARS-CoV-2 N1 or N2 genes, they needed to average the manufacturer’s limit of detection of >/= 0.1 copies/µL and 2 positive droplets per well.

### SALIVARY MEASUREMENT OF ANTIBODIES AGAINST THE SARS-COV-2 PROTEINS

The luciferase immunoprecipitation systems (LIPS) immunoassay was used to study IgG antibody response against SARS-CoV-2 in saliva. Due to the potential biohazard of infectious SARS-CoV-2 in saliva and a viral inactivation protocol involving heating saliva at 56 C o for 30 min was employed. For these SARS-CoV-2 antibody measurements, Renilla luciferase-nucleocapsid and Gaussia luciferase-spike fusion protein extracts were employed with protein A/G beads as the IgG capture reagent as previously described (72). Due to the lower levels of immunoglobulin present in saliva, 10 uL of each saliva sample was utilized in the LIPS assay as previously described (73). Known SARS-CoV-2 serum samples for IgG antibodies against nucleocapsid and spike proteins and saliva from uninfected controls were used for assigning seropositive cut-off values and for standardization.

## Data Availability

DATA AVAILABILITY:
Unpublished human oral single cell datasets (minor salivary glands and oral mucosa) in this
study can be visualized and assessed at www.covid19cellatlas.org. The published oral datasets from mice can accessed via the Chan Zuckerberg Biohub at https://tabulamuris.ds.czbiohub.org/ (dorsal tongue47) or via the Gene Expression Omnibus (https://www.ncbi.nlm.nih.gov/geo/) under the accession numbers GSE113466 (submandibular
glands67), GSE131285 (parotid glands), and GSE120654 (buccal mucosa46). Human neutrophil
data can be downloaded using the SRA Toolkit (SRP27137550)
CODE AVAILABILITY:
Analysis notebooks are available at github.com/Teichlab/covid19_oral.

https://tabulamuris.ds.czbiohub.org/

https://www.ncbi.nlm.nih.gov/geo/

https://github.com/Teichlab/covid19_oral

https://www.humancellatlas.org/publications/

http://software.10xgenomics.com/single-cell/overview/welcome

https://satijalab.org/

## DATA AVAILABILITY

Unpublished human oral single cell datasets (minor salivary glands and oral mucosa) in this study can be visualized and assessed at www.covid19cellatlas.org. The published oral datasets from mice can accessed via the Chan Zuckerberg Biohub at https://tabula-muris.ds.czbiohub.org/ (dorsal tongue47) or via the Gene Expression Omnibus (https://www.ncbi.nlm.nih.gov/geo/) under the accession numbers GSE113466 (submandibular glands67), GSE131285 (parotid glands), and GSE120654 (buccal mucosa46). Human neutrophil data can be downloaded using the SRA Toolkit (SRP27137550).

## CODE AVAILABILITY

Analysis notebooks are available at github.com/Teichlab/covid19_oral.

## Acknowledgements

This work is dedicated to the more than 1 million worldwide fatalities and their families who have fallen victim to COVID-19. These experiments were principally supported by grants and awards to K.M.B. (NIDCR K08 DE026537; AAP Sunstar Innovation Grant; L30 Loan Repayment Program for Clinical Researchers), and to B.M.W. (NIDCR Z01-DE000704) through the National Institutes of Health, National Institute of Dental and Craniofacial Research, Division of Intramural Research. Gingival scRNAseq was conducted by the UNCAdvanced Analytics Core (Center for GI Biology and Disease; P30 DK034987). Additional support is from the Senior Research Training Fellowship by the American Lung Association (RT-575362), the Postdoctoral Research Fellowship by the Cystic Fibrosis Foundation (KATO20F0) to T.K, and NIH/NIDCR (K01DE027087 to J.T.M). The following investigators are also supported by the NIH Intramural Research Program: J.A.C., L.A.T., D.E.K., S.H., D.C., S.P., J.L. All patients seen at the author’s (B.M.W.) institute (NIDCR) reported herein provided informed consent prior to participation in IRB-approved research protocols (NIH IRB: 20-D-0094, NCT04348240; NIH IRB: 15-D-0051, NCT02327884). A special acknowledgement to the NIH Office of the Director and the NIH Principal Deputy Director, Lawrence A. Tabak, for his profound assistance in establishing the NIH COVID-19 Saliva Testing Study. We would like to acknowledge the NIDCR Office of the Clinical Director and the Office of the Scientific Director for their COVID-19-related support including assistance in protocol development and research activity support. And also, special thanks to Ms. Beth Brillante and Ms. Caroline Webb for their assistance in protocol navigation and project management. Healthy volunteer and COVID-19 clinical specimens were collected by members of NIDCR Sjgren’s Syndrome Clinic. A special thanks for the coordinating efforts of the NIH COVID-19 Testing Facility, the NIH Occupational Medicine Service, and NIH Medical Records for their support in executing the NIH Clinical Study. scRNAseq and bulk RNAseq of the salivary glands were conducted by the NIDCR Genomics and Computational Biology Core and a very deserving thanks to Drs. Zheng Wei and Dani Martin for their assistance in executing and processing the salivary gland scRNAseq captures. Mr. Willie Young for his assistance in performing the COVID-19 autopsies. This publication is part of the Human Cell Atlas - www.humancellatlas.org/publications.

## Author Contributions

Conceptualization: RCB, SAT, JL, JC, BMW & KMB

Methodology: NH, GC, CA, MF, RCB, KF, JL, LT, PB, BMW & KMB

Patient Recruitment and Data Collection: EP, MB, DC, BMW

Biospecimen Collection: RM, SW, MW, RP, AK, VM, JM, MB, BG, SS, DC, SH, DK, SP, MB, JM, KMB, RM, BMW

Autopsy Tissue and Data Collection: NIH COVID-19 Autopsy Consortium Bioinformatics: NH, CDC, TP, BF

Investigation & Data Analysis: NH, PP, TK, YM, KO, CDC, KF, MF, PB, BMW & KMB

Writing – Original Draft: KMB & BMW

Writing – Review & Editing: NH, CDC, TK, YM, PP, PB, RB, SAT, BMW & KMB

## Declaration of Interests

The authors declare that they have no conflict of interest. The authors had access to the study data and reviewed and approved the final manuscript.

**GROUP AUTHORSHIPS:**

**HCA ORAL & CRANIOFACIAL BIOLOGICAL NETWORK**

Kevin M. Byrd^1,2^, Ins Sequeira^3^, Blake M. Warner^4,5^, Sarah Teichmann^6,7^, Marcelo Freire^8,9^, Adam Kimple^10^

1 Division of Oral & Craniofacial Health Sciences, University of North Carolina Adams School of Dentistry, Chapel Hill, NC, USA; 2 ADA Science & Research Institute, Gaithersburg, MD, USA; 3 Centre for Oral Immunobiology and Regenerative Medicine, Barts and The London School of Medicine and Dentistry, Queen Mary University of London; 4 Salivary Disorders Unit, National Institute of Dental and Craniofacial Research, National Institutes of Health, Bethesda, Maryland, USA; 5 Sjgren’s Syndrome Clinic, National Institute of Dental and Craniofacial Research, National Institutes of Health, Bethesda, Maryland, USA; 6 Wellcome Sanger Institute, Wellcome Genome Campus, Hinxton, Cambridge CB10 1SA, UK; 7 Dept Physics, Cavendish Laboratory, JJ Thomson Ave, Cambridge CB3 0HE, UK; 8 Department of Genomic Medicine, J. Craig Venter Institute, La Jolla, California, USA; 9 Department of Infectious Disease, J. Craig Venter Institute, La Jolla, California, USA; 10 Department of Otolaryngology-Head and Neck Surgery, University of North Carolina School of Medicine Chapel Hill, NC, USA

**NIH COVID-19 AUTOPSY CONSORTIUM**

**National Institutes of Health, Bethesda, Maryland, USA**

*Laboratory of Pathology, Center for Cancer Research, National Cancer Institute*

David E. Kleiner MD, PhD; Billel Gasmi MD; Michelly Sampaio De Melo MD; Esra Dikoglu, MD; Sabina Desar, MD; Stephen M Hewitt, M.D., Ph.D.; Kris Ylaya; Joon-Yong Chung, Ph.D.; Stefania Pittaluga MD, PhD; Grace Smith

*Critical Care Medicine Department, NIH Clinical Center*

Daniel S. Chertow, MD, MPH; Kevin M. Vannella, PhD; Sydney Stein, DVM; Marcos Ramos-Benitez, PhD; Sabrina C. Ramelli, PhD; Shelly J. Curran, PhD; Ashley L. Babyak; Luis Valenica Perez; Mary E. Richert, MD

*Kidney Diseases Branch, National Institute of Diabetes and Digestive and Kidney Diseases*

Alison Grazioli, MD

**University of Maryland Baltimore, Maryland, USA**

*University of Maryland Baltimore School of Medicine*

Nicole Hays; Madeleine Purcell; Shreya Singireddy; Jocelyn Wu; Jean Chung, CRNP; Amy Borth, MSN, ACNP; Kimberly Bowers, ACNP-BC; Anne Weichold, CRNP

*Department of Surgery, Division of Cardiac Surgery*

Douglas Tran, MD; Ronson J. Madathil, MD

*Department of Surgery, Division of Thoracic Surgery*

Eric M. Krause, MD, JD

*Department of Shock Trauma Critical Care*

Daniel L. Herr, MD

*Department of Surgery, R Adams Cowley Shock Trauma Center*

Joseph Rabin, MD; Joseph A. Herrold, MD, MPH; Ali Tabatabai, MD; Justin E. Richards, MD; Erich Hochberg, MPAS, PA-C; Christopher Cornachione, MSN, CRNP

*Department of Medicine, Division Pulmonary & Critical Care Medicine*

Andrea R. Levine, MD; Michael T. McCurdy, MD; Mary E. Richert, MD

*Department of Medicine, Division of Infectious Disease*

Kapil K. Saharia, MD, MPH

*Department of Anesthesiology, Division of Critical Care Medicine*

Zack Chancer, MD, MS; Michael A. Mazzeffi, MD; Justin E. Richards, MD

**University of Maryland, St. Joseph Medical Center, Towson, Maryland, USA**

*Department of Pathology*

James W. Eagan, Jr, MD

*Tidal Health Peninsula Regional, Department of Pulmonology*,

Yashvir Sangwan, MBBS

---

**Supp. Figure 1.**
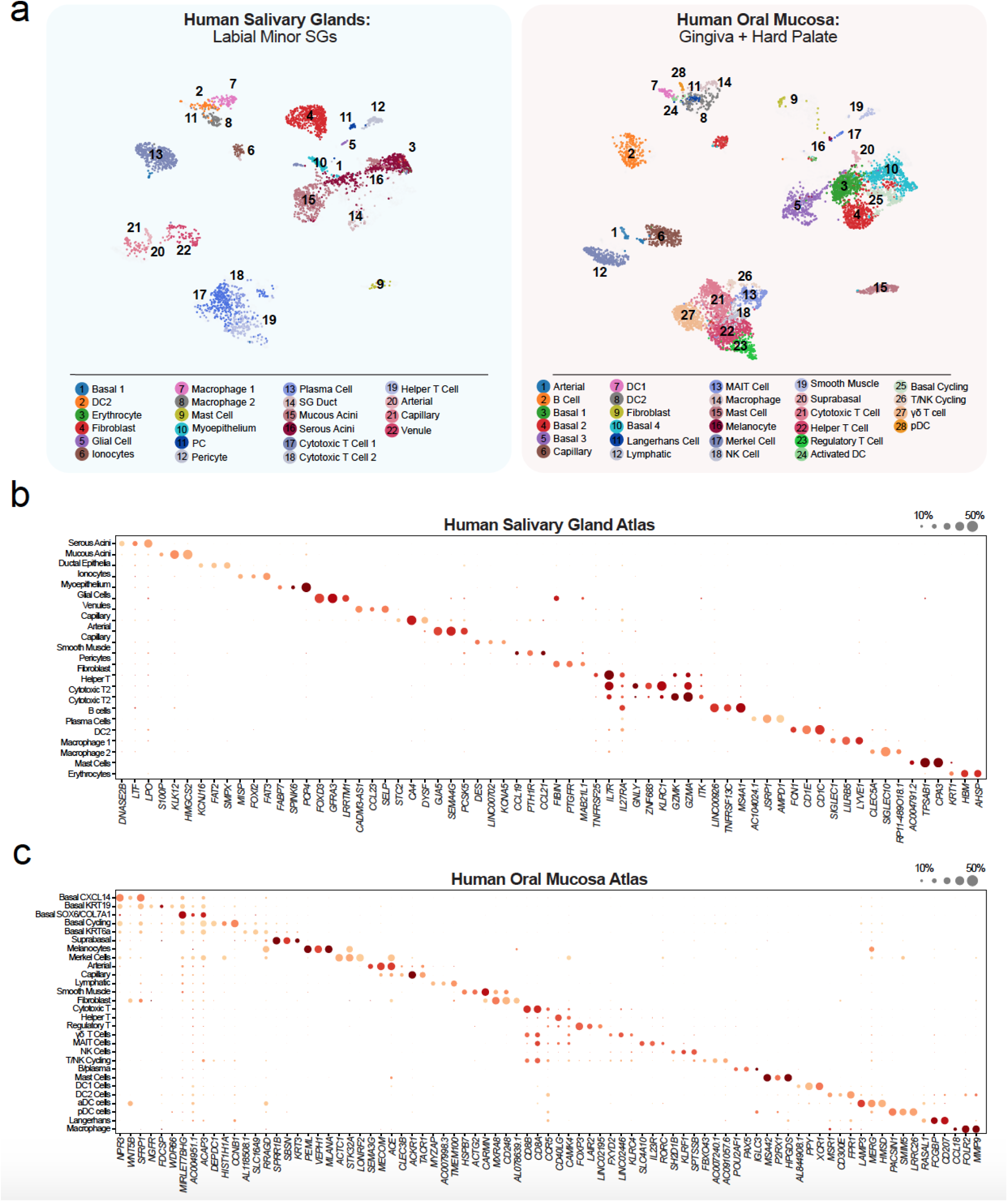
Distinct oral cell type signatures are found in human oral tissue niches. **(a)** Using unpublished data sets, UMAPs of single cell RNA sequencing data from dissociated gingiva and minor saliva glands delineates cell population contribution by sample and cell type annotation. We found 22 populations in the glands (3370 cells included) and 28 populations in the gingiva (5340 cells). Labial minor SGs were procured from patients having biopsies for the clinical workup of Sjögren’s Syndrome who did not meet classification criteria and were otherwise healthy, including lacking SSA/SSB autoantibodies, and without focal lymphocytic sialadenitis. Mucosal tissues including the gingiva and hard palate were harvested from four 20 to 30-year-old healthy subjects who were given a stent that prevented brushing only on three maxillary teeth to induce a very localized gingivitis (65, 66). **(b)** The top three defining minor SG atlas genes expressed in each annotated cluster reveal unique epithelial and immune cell heterogeneity. This includes 5 epithelia cell populations and (2 macrophage populations, 2 cytotoxic T cell populations). These annotations serve as niche atlases but also the reference for the human pan-oral atlas in Figure 2. **(c)** The top three defining gingival atlas genes expressed in each annotated cluster reveal significant epithelial and immune cell heterogeneity. This includes at least 7 populations of oral epithelial basal cells (Basal 1-4, Basal cycling; Merkel Cells, and Melanocytes) and 15 immune cell subsets (3 DC subsets, 7 T cell subsets).

**Supp. Figure 2.**
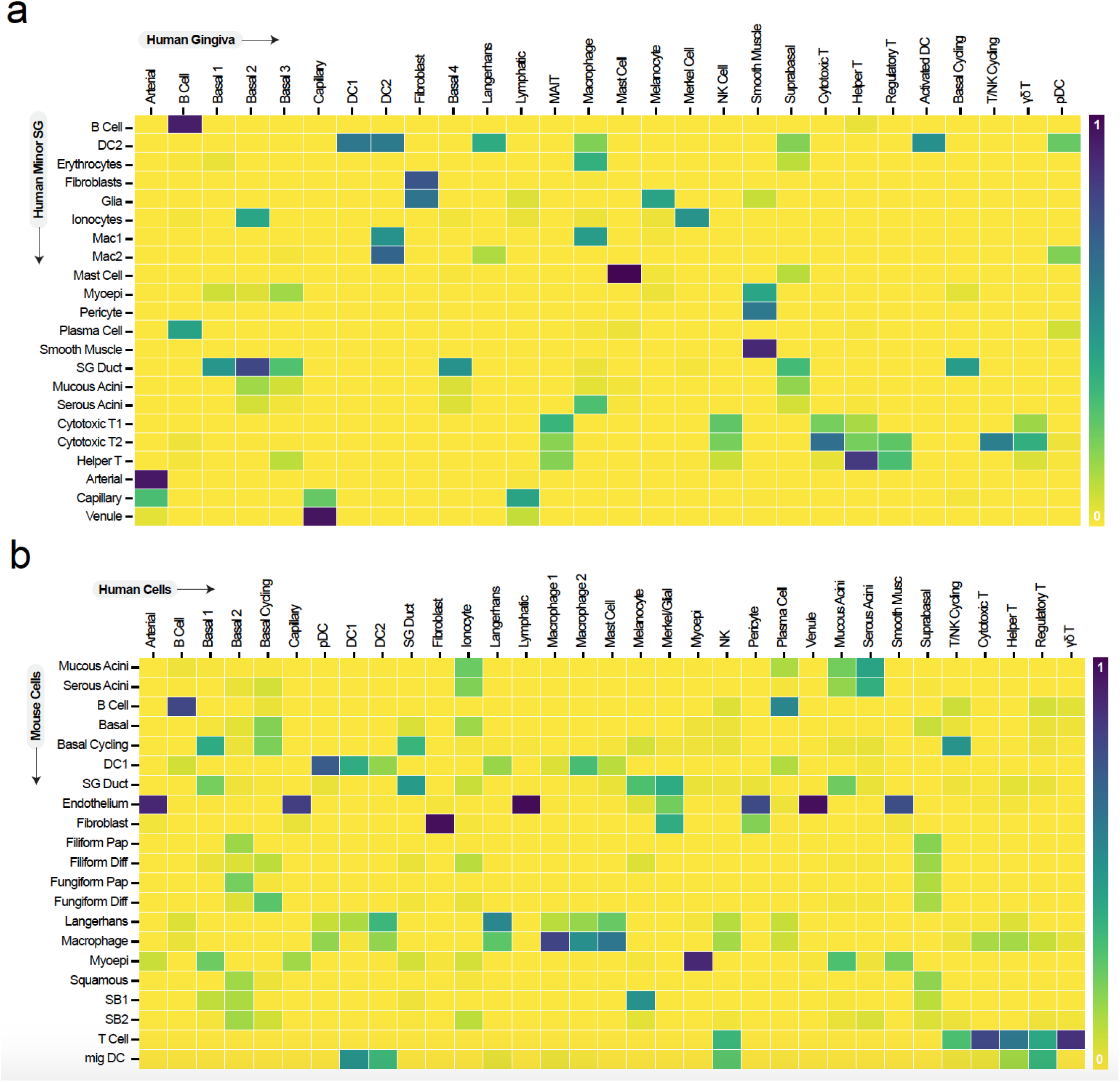
Cluster correlation analyses of oral atlases supports further exploration. **(a)** Correlation mapping between human gingival and saliva gland cell clusters (see Supp. Figure 1) reveal expectedly invariant clusters are well matched between samples (B cells, fibroblasts, mast cells, smooth muscle, helper T cells, arterial cells). However, numerous other cell annotation heterogeneities remain to be further explored (Mac1, Mac2 in saliva glands; Basal 1-4 in gingiva). **(b)** Similar correlation mapping between mouse and human pan-oral joint annotations support similarities and unique differences between cell clusters between species. B cells again are well correlated as are fibroblasts, myoepithelium, and macrophages. However, the overlap in endothelial populations from mice into arterial, capillary, lymphatic, pericyte, venule, and smooth muscle human clusters suggests more cells are suggests that either further exploration of the mouse oral blood vessel populations at higher resolution, or that universal nomenclature of cell types is needed. Additionally, other matched scRNAseq sites in humans (adding dorsal tongue, buccal mucosa, tonsil, major saliva glands) and mice (adding gingiva and hard palate) will also better relate newly discovered populations.

**Supp. Figure 3.**
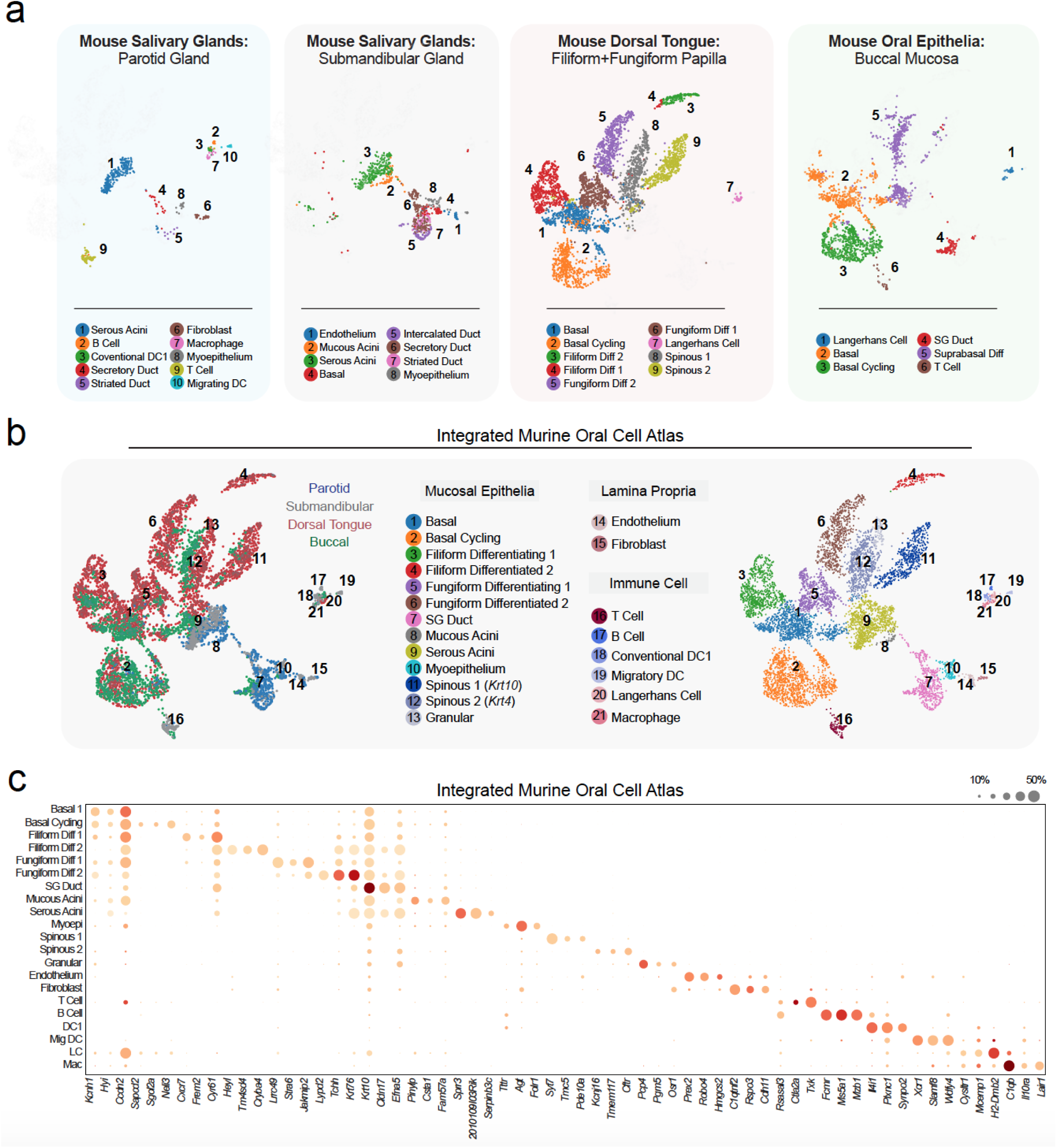
An integrated murine pan-oral cell atlas supports cell heterogeneity. **(a**,**b)** Individual single cell datasets from mice were normalized and reannotated before integration. We found 10 populations in the parotid gland (438 cells included), 8 populations in the submandibular gland (763 cells), 9 populations in the dorsal tongue (3610 cells), and 6 populations (1661 cells), some of which were not originally reported. These 33 populations were jointly annotated to represent 21 populations in the pan-oral atlas. (b) Using published data sets (46-48, 67), we integrated across available single cell RNA sequencing datasets to describe the cell types vulnerabilities in mouse oral cavity tissues. (Left) UMAPs of dorsal tongue, buccal mucosa, submandibular gland, and parotid gland by color highlights tissue contribution. (Right) Joint annotation finds both shared and unique cell types (e.g., filiform and fungiform epithelia) across expected tissue compartments (epithelia, lamina propria, and immune cell populations). **(c)** Dot plot expression matrix visualization of the integrated mouse oral atlas shows that cell types are distinguished by unique transcripts, some of which are previously undescribed.

**Supp. Figure 4.**
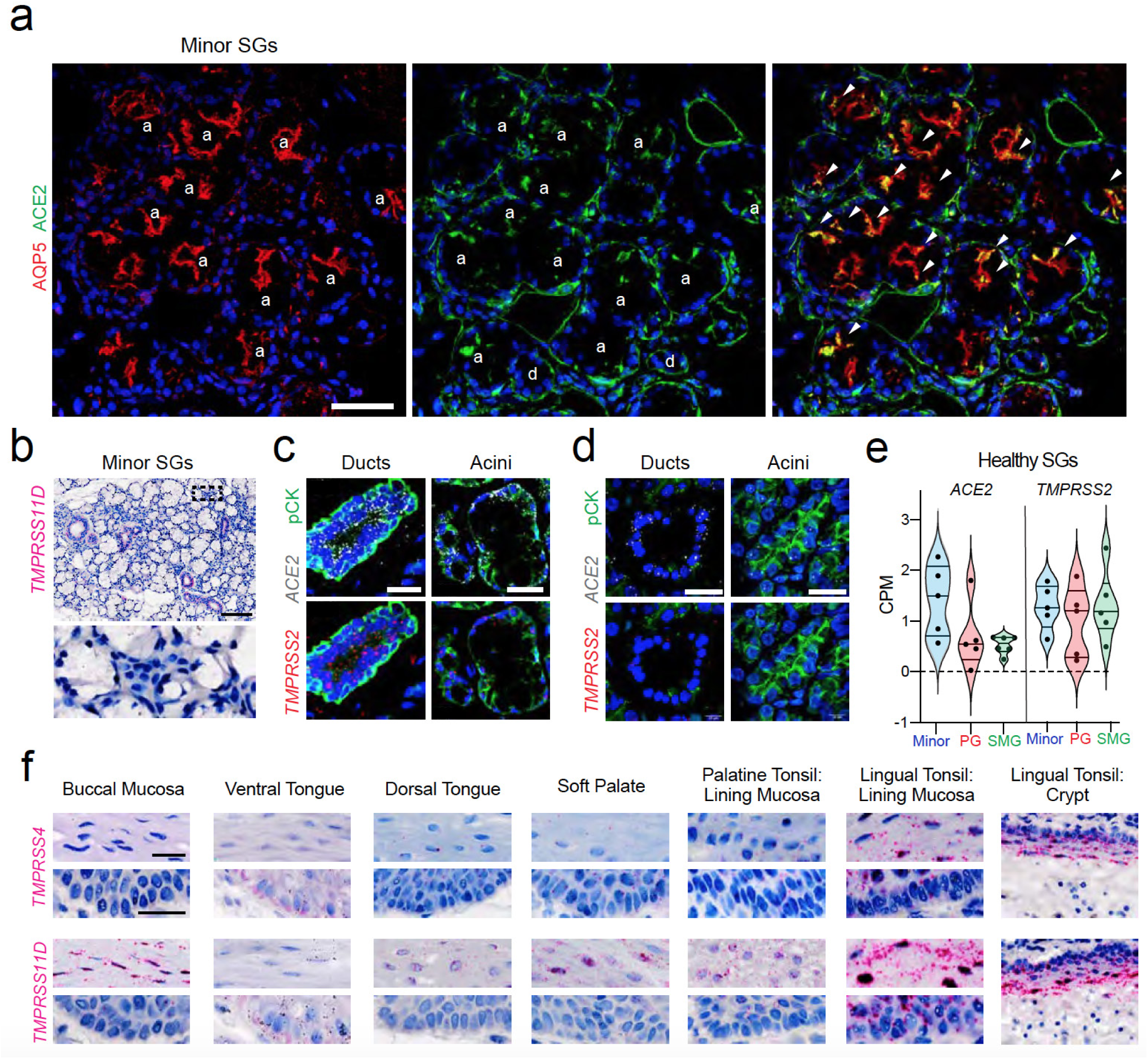
Validation of SARS-CoV-2 expression patterning across oral niches. **(a)** Immunofluorescent (IF) confocal microscopy demonstrated AQP5 (red, acini) colocalization with ACE2 (green) reveal that *ACE2* is enriched on the apical (luminal) and basolateral membranes and is concentrated in the SG acini/ducts (labeled ‘a’ or ‘d’). White arrows indicate colocalization between *ACE2* and AQP5. *ACE2* is also expressed to a similar degree in the saliva gland ducts but rarely expressed in myoepithelial cells circumscribing the acini (data not shown). **(b-d) (b)** Using healthy volunteer gland sections, the unique minimal *TMPRSS11D* expression in ducts and acini was confirmed using RNAscope® in situ hybridization. **(c**,**d)** To further confirm co-expression of *ACE2* and *TMPRSS2*, RNAscope® fluorescent in situ hybridization and immunohistochemistry for pan-cytokeratin (pCK), shows that acini and ducts co-express *ACE2* and *TMPRSS2* further highlighting their vulnerability to infection in (c) minor and (d) parotid saliva glands. **(e)** Analysis of available bulk RNA sequencing data suggests that the minor (minor) and parotid (PG) saliva glands are vulnerable to SARS-CoV-2 infection compared to the submandibular glands (SMG). The minor glands express ∼3x higher *ACE2* compared to the major glands. For comparison, the gland express an equivalent amount of TMPRSS2 across all three glands samples. **(f)** Considering the unique niches of the oral cavity, we used ISH mapping for *ACE2* and *TMPRSS2* (for *ACE2* and *TMPRSS2*, see Figure 4) across the oral (buccal mucosa, ventral tongue, and the dorsal tongue) as well as examining the oropharynx for the first time (soft palate, tonsils). This again supports oral niche heterogeneity and that all sites are vulnerable to infection in suprabasal cells that are sloughed into saliva. Dotted black box in (b) represents zoomed-in area. Scale bars: (**b**) 100μm, (**a**) 50μm, (**c**,**d**,**f**) 25μm.

**Supp. Figure 5.**
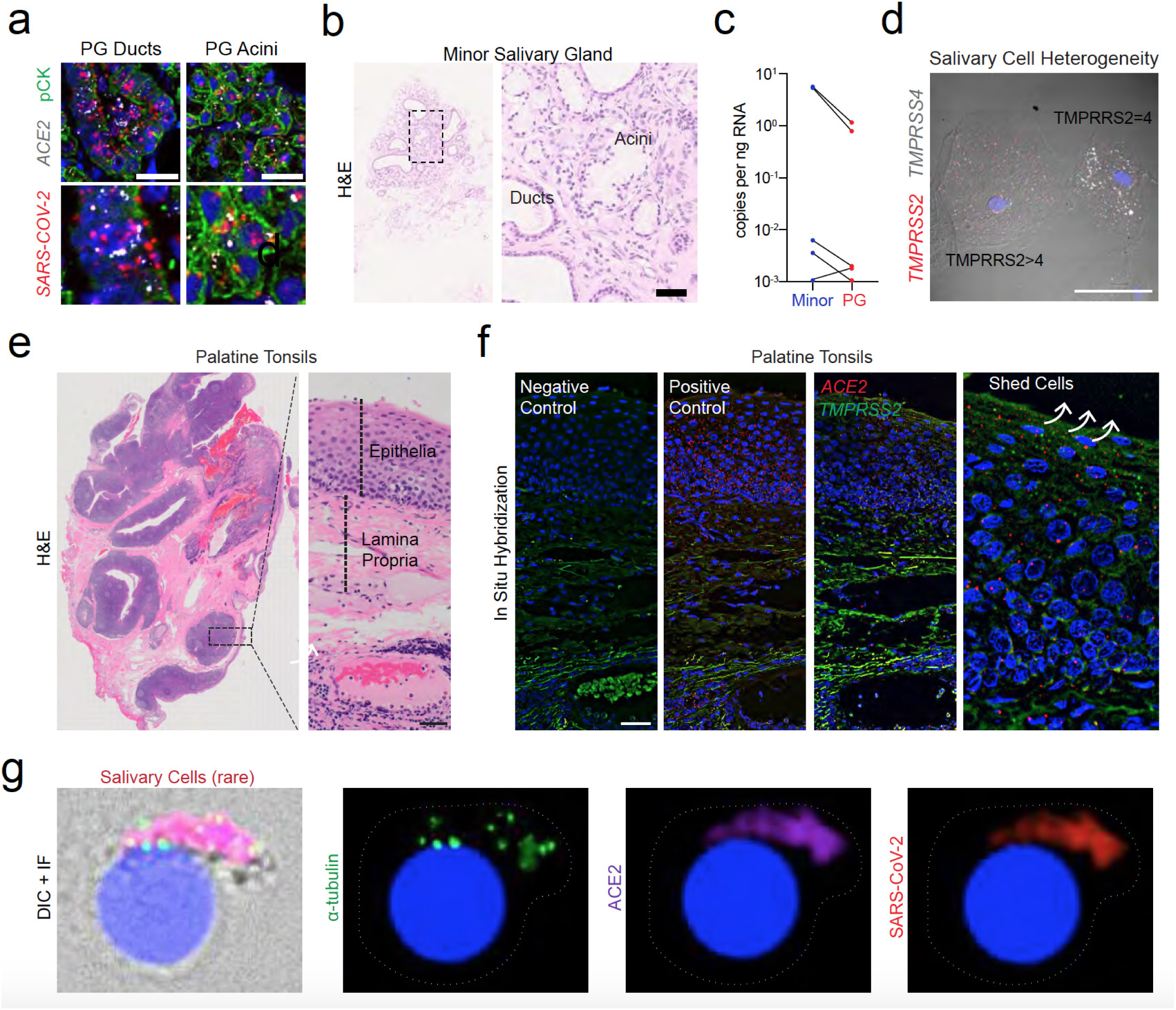
An oral infection axis occurs via glands and suprabasal epithelia. **(a)** In situ hybridization (ISH) also reveals infection in the major parotid gland (PG).**(b)** Microscopic assessment of the COVID-19 saliva glands by H&E reveals variable histological presentations including atrophy and fibrosis. **(c)** Viral load in minor saliva glands is generally higher when compared to glands using ddPCR. **(d)** Shed cells in saliva display unique TMPRSS family heterogeneity, which we also observed in tissue-specific oral atlases for glands and mucosa. Saliva glands express the highest TMPRSS2 in mouse and human atlases, and some shed epithelial cells express similar patterns, suggesting they may be shed from the SG ducts. Others, express higher TMPRSS4, suggesting these cells may be from the suprabasal mucosa. **(e-f) (e)** ISH mapping validation pipeline for discovering epithelial expression of SARS-CoV-2 entry factor expression in shedding suprabasal cells, first starting with H&E histochemical staining. **(f)** ISH was used with negative and positive probe controls. **(g)** Ciliated cells in saliva are exceedingly rare (α-tubulin+) but can present and infected. Dotted black box in **(b**,**e)** represents zoom-in regions; white arrow in **(f)** represents sloughing trajectory; dotted white lines **(g)** highlight cell membranes; arrowheads in **(g**,**h)** indicate SARS-CoV-2+ (red) cells. Scale bars: **(b)** 50μm **(a**,**d**,**e**,**h)** 25μm, **(g)** 10μm.

**Supp. Figure 6.**
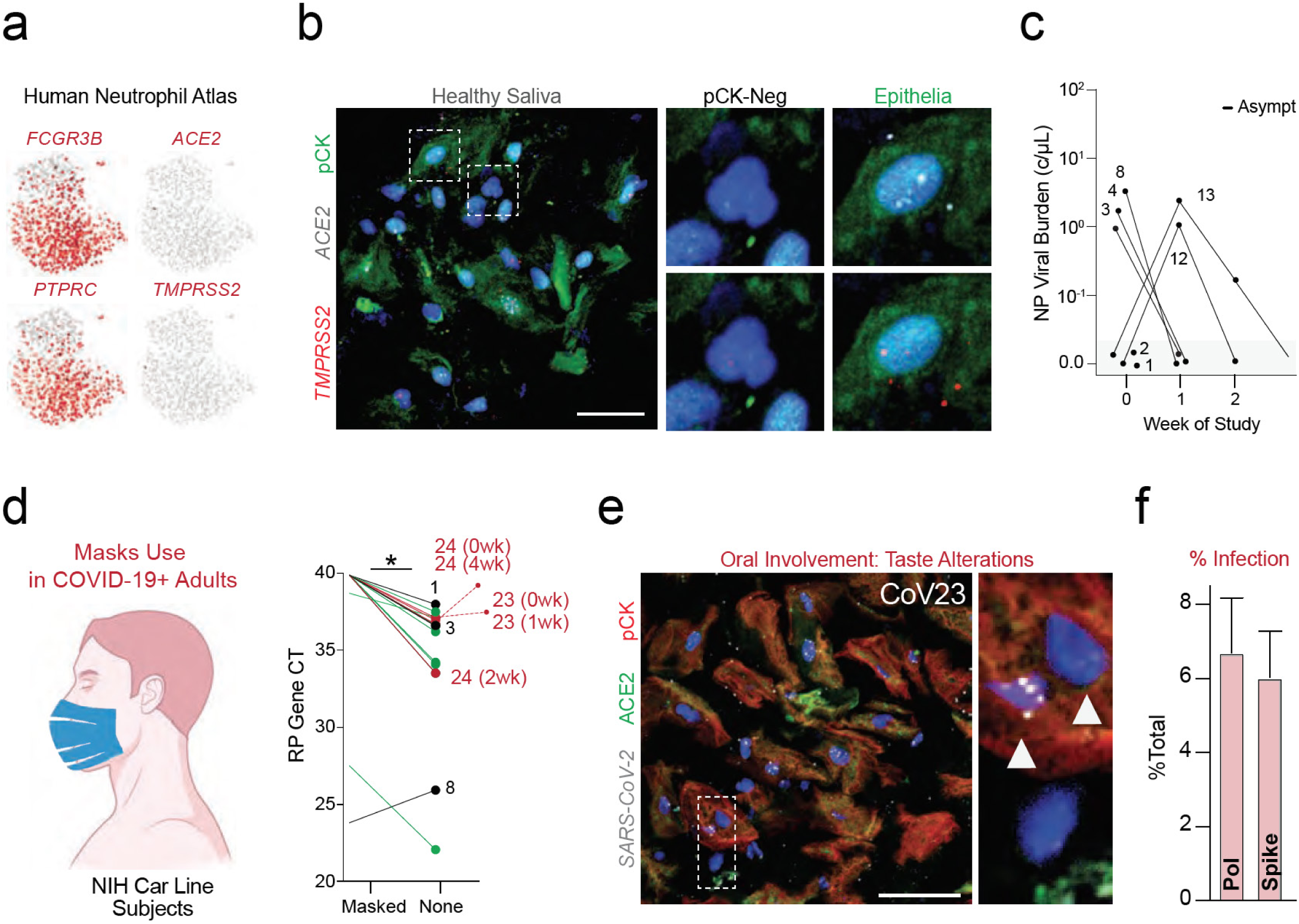
Prospective clinical sampling confirms epithelial cell infection. **(a)** Using a recently pre-printed salivary neutrophil atlas from healthy patients50, we confirm that virtually no neutrophils express SARS-CoV-2 *ACE2* or *TMPRSS2*. Neutrophils constitute ∼50% of salivary cells and epithelial cells the other 50% with trace amounts of lymphocytes and other cell populations. **(b)** Using cell blocks of heathy saliva, we validate our findings that shed salivary epithelial (pan-cytokeratin positive; pCK+) cells express both *ACE2* and *TMPRSS2*. We see minimal to no expression of these SARS-CoV-2 entry factors in pCK-negative cells. **(c)** From the NIH Car Line study, the majority of asymptomatic subjects only displayed NP swab positivity, including two patients who developed symptoms during the prospective sampling (CoV12 and CoV13). **(d)** Taking this cohort of asymptomatic and symptomatic subjects, infectious saliva fraction was measured with masks and unmasked highlighting the need for masks to prevent the spread of SARS-CoV-2 via saliva droplets. While obvious, this measure highlights the ability to prevent the spread of the oral infection axis (see Cov01 saliva ejection with no mask ‘none’). **(e**,**f) (e)** We confirm our findings from the UNC OBS-C study and from NIH Car Line study that highest saliva viral load and reported taste alterations (dysgeusia in CoV19), we observe significant salivary epithelial (pCK+)cells; See also Figure 6h. (f) results of pol and spike quantification of salivary cell infection among 3 samples show consistency of the infection quantification method. Scale bars: **(a**,**e)** 25μm.

**Supplemental Table 1.**
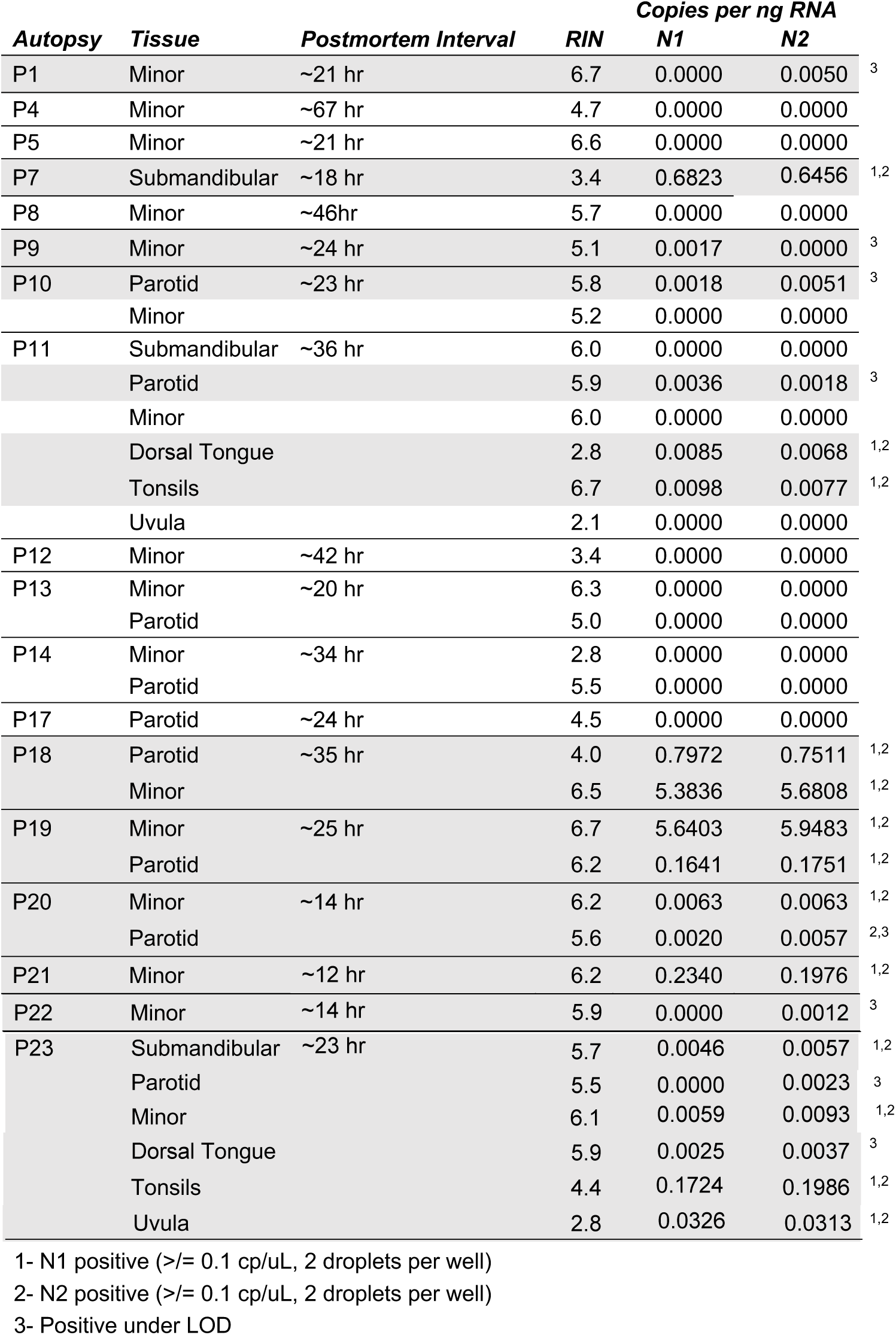
Tissue types, specimen characteristics, and droplet digital PCR results for CDC SARS-CoV-2 testing.

| Autopsy | Tissue | Postmortem Interval | RIN | Copies per ng RNA |  |  |
| --- | --- | --- | --- | --- | --- | --- |
|  |  |  |  | N1 | N2 |  |
| P1 | Minor | ~21 hr | 6.7 | 0.0000 | 0.0050 | 3 |
| P4 | Minor | ~67 hr | 4.7 | 0.0000 | 0.0000 |  |
| P5 | Minor | ~21 hr | 6.6 | 0.0000 | 0.0000 |  |
| P7 | Submandibular | ~18 hr | 3.4 | 0.6823 | 0.6456 | 1,2 |
| P8 | Minor | ~46hr | 5.7 | 0.0000 | 0.0000 |  |
| P9 | Minor | ~24 hr | 5.1 | 0.0017 | 0.0000 | 3 |
| P10 | Parotid | ~23 hr | 5.8 | 0.0018 | 0.0051 | 3 |
|  | Minor |  | 5.2 | 0.0000 | 0.0000 |  |
| P11 | Submandibular | ~36 hr | 6.0 | 0.0000 | 0.0000 |  |
|  | Parotid |  | 5.9 | 0.0036 | 0.0018 | 3 |
|  | Minor |  | 6.0 | 0.0000 | 0.0000 |  |
|  | Dorsal Tongue |  | 2.8 | 0.0085 | 0.0068 | 1,2 |
|  | Tonsils |  | 6.7 | 0.0098 | 0.0077 | 1,2 |
|  | Uvula |  | 2.1 | 0.0000 | 0.0000 |  |
| P12 | Minor | ~42 hr | 3.4 | 0.0000 | 0.0000 |  |
| P13 | Minor | ~20 hr | 6.3 | 0.0000 | 0.0000 |  |
|  | Parotid |  | 5.0 | 0.0000 | 0.0000 |  |
| P14 | Minor | ~34 hr | 2.8 | 0.0000 | 0.0000 |  |
|  | Parotid |  | 5.5 | 0.0000 | 0.0000 |  |
| P17 | Parotid | ~24 hr | 4.5 | 0.0000 | 0.0000 |  |
| P18 | Parotid | ~35 hr | 4.0 | 0.7972 | 0.7511 | 1,2 |
|  | Minor |  | 6.5 | 5.3836 | 5.6808 | 1,2 |
| P19 | Minor | ~25 hr | 6.7 | 5.6403 | 5.9483 | 1,2 |
|  | Parotid |  | 6.2 | 0.1641 | 0.1751 | 1,2 |
| P20 | Minor | ~14 hr | 6.2 | 0.0063 | 0.0063 | 1,2 |
|  | Parotid |  | 5.6 | 0.0020 | 0.0057 | 2,3 |
| P21 | Minor | ~12 hr | 6.2 | 0.2340 | 0.1976 | 1,2 |
| P22 | Minor | ~14 hr | 5.9 | 0.0000 | 0.0012 | 3 |
| P23 | Submandibular | ~23 hr | 5.7 | 0.0046 | 0.0057 | 1,2 |
|  | Parotid |  | 5.5 | 0.0000 | 0.0023 | 3 |
|  | Minor |  | 6.1 | 0.0059 | 0.0093 | 1,2 |
|  | Dorsal Tongue |  | 5.9 | 0.0025 | 0.0037 | 3 |
|  | Tonsils |  | 4.4 | 0.1724 | 0.1986 | 1,2 |
|  | Uvula |  | 2.8 | 0.0326 | 0.0313 | 1,2 |

1- N1 positive ( $\geq 0.1$ cp/uL, 2 droplets per well)
2- N2 positive ( $\geq 0.1$ cp/uL, 2 droplets per well)
3- Positive under LOD

**Supplemental Table 2.**
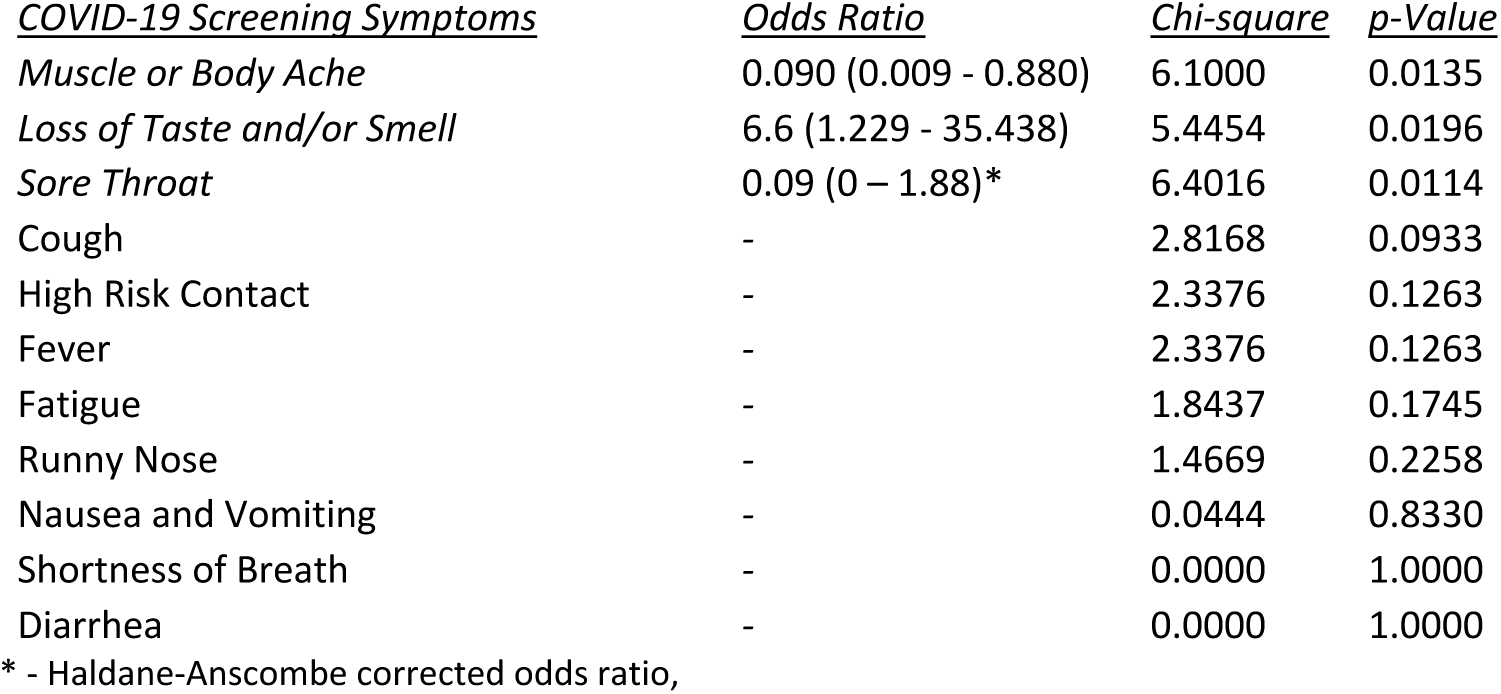
COVID-19 Symptoms correlate with oral cavity infections.

| <u>COVID-19 Screening Symptoms</u> | <u>Odds Ratio</u> | <u>Chi-square</u> | <u>p-Value</u> |
| --- | --- | --- | --- |
| *Muscle or Body Ache* | 0.090 (0.009 - 0.880) | 6.1000 | 0.0135 |
| *Loss of Taste and/or Smell* | 6.6 (1.229 - 35.438) | 5.4454 | 0.0196 |
| *Sore Throat* | 0.09 (0 – 1.88)* | 6.4016 | 0.0114 |
| Cough | - | 2.8168 | 0.0933 |
| High Risk Contact | - | 2.3376 | 0.1263 |
| Fever | - | 2.3376 | 0.1263 |
| Fatigue | - | 1.8437 | 0.1745 |
| Runny Nose | - | 1.4669 | 0.2258 |
| Nausea and Vomiting | - | 0.0444 | 0.8330 |
| Shortness of Breath | - | 0.0000 | 1.0000 |
| Diarrhea | - | 0.0000 | 1.0000 |

\* - Haldane-Anscombe corrected odds ratio,

## Notes

### Competing Interest Statement

In the last year KMB has been a Scientific Advisor at Arcato Laboratories, Inc. and Coordinator at the Human Cell Atlas. For the last month MOF has served as a Scientific Advisor for PolyBio Research Foundation. In the last 3 years SAT has been remunerated for consulting by Roche and Genentech, and is a member of Scientific Advisory Boards at Biogen, GlaxoSmithKline and Foresite Labs.

### Clinical Trial

NCT04348240; NCT02327884; NCT04105569

### Author Declarations

The studies were conducted at two sites, independently utilizing clinical protocols which were approved by institutional review boards (IRB) at the respect institutions. UNC: Single cell gingival atlas constructed from human tissues was registered Clinicaltrials.gov ID: NCT04105569, approved by UNC IRB: 19-0183. Participant tissues included remnant oral specimens previously collected through the University of North Carolina Adams School of Dentistry Oral Pathology Biobank (UNC IRB 20-1501) or the Center for Oral and Systemic Diseases (COSD) Biorepository (UNC IRB 15-1814). Lab and project personnel were double-blinded and did not have access to PHI or any links to any PHI. Project members did not have interpersonal contact with patients that provided specimens. A subset of 10 samples from COVID-19 outpatients were collected (approved by UNC IRB 20-0792). The purpose of this study is to describe the epidemiology, clinical features, and immunological response to SARS-CoV-2 infection and develop new diagnostic tests focused on saliva. NIH: All patients seen at the author (B.M.W.) institute (NIDCR) reported herein provided informed consent prior to participation in NIH IRB-approved research protocols: NIH IRB 20-D-0094, NCT04348240; NIH IRB 15-D-0051, NCT02327884.

